# Association of neighborhood deprivation with Alzheimer’s Disease pathology, brain structure, and cognition by race and ethnicity, sex, and APOE ε4 status

**DOI:** 10.64898/2026.05.05.26352370

**Authors:** Pablo Aguilar-Dominguez, Carmen-Maria Colceriu, Thomas Monroe Holland, Samuel N Lockhart, Joseph C. Masdeu, Lycia Tramujas Vasconcellos Neumann, Heather M Snyder, Laura D Baker, Alexandre Bejanin, Susan M. Landau, Eider Arenaza-Urquijo

## Abstract

**BACKGROUND:** We investigated associations of neighborhood disadvantage with Alzheimer’s Disease (AD)-related outcomes by biological and social factors in at-risk older adults.

**METHODS:** 1,880 U.S. POINTER participants with Area Deprivation Index (ADI) and cognition (PACC) were included. 868 had amyloid, tau PET, white matter hyperintensities (WMH), and/or gray matter volumes. We conducted exploratory, linear models testing ADI interactions with sex, race and ethnicity, and APOE ε4, adjusting for age and education.

**RESULTS:** “White/European American”, “Hispanic/Latinx/Spanish” and “Others” showed lower cognitive scores with higher ADI, while “White/European American” showed the highest cognitive scores across ADI levels. APOE ε4 carriers from high-ADI areas showed higher WMH and tau, and “Hispanic/Latinx/Spanish” from more deprived areas showed higher WMH. Females from moderate-ADI areas showed higher tau. Amyloid burden was higher in APOE ε4 carriers from low-ADI areas.

**CONCLUSION:** Differential associations of ADI with AD-related outcomes across biological and social factors may reflect systemic health disparities and study design.

## 1 | Background

More than 50 million people worldwide are living with dementia^1^, with Alzheimer’s disease (AD) being the leading cause^2^. The prevalence of dementia is expected to double within the next two decades^3^, representing a major global health and social challenge. Lifelong exposure to adverse structural and social determinants of health (SDOH) and associated inequities negatively affect brain health and may increase susceptibility to clinical Alzheimer’s disease and Alzheimer’s disease–related dementias (AD/ADRD)^4^. Elucidating the pathways by which SDOH contribute to dementia risk is therefore essential.

Neighborhood-level socioeconomic disadvantage –a key SDOH factor– has been associated with higher dementia incidence and cognitive impairment^5,6^. Further, epidemiological research suggests that neighborhood disadvantage may intersect with individual-level factors such as sex, gender, race or ethnicity to increase dementia risk. Thus, women residing in socioeconomically deprived neighborhoods are at higher risk of developing dementia and AD compared to men^7^. Further, neighborhood disadvantage may account for a substantial proportion of the elevated risk for ADRD in Non-Hispanic Black and Hispanic older adults relative to Non-Hispanic White older adults^8^.

The biological mechanisms underlying the association of neighborhood-level socioeconomic disadvantage with dementia risk, particularly as they intersect with individual-level factors, remain insufficiently characterized. A commonly used indicator of neighborhood disadvantage is the Area Deprivation Index (ADI), which ranks neighborhoods at the Census Block Group level^9,10^ based on income, education, employment, and housing quality^9^, with higher ADI values indicating greater neighborhood-level socioeconomic disadvantage. A limited number of studies have examined the association of neighborhood disadvantage, via ADI or other proxies, with AD-related outcomes. Among cognitively unimpaired older adults, studies suggest that higher neighborhood deprivation is associated with multiple AD-related outcomes, including lower global cognition^11^, lower hippocampal brain volume^12,13^, higher total tau measured with cerebrospinal fluid (CSF)^14^, and greater post-mortem amyloid burden^15^. Evidence linking neighborhood deprivation and white matter hyperintensities (WMH) is limited and mixed: some studies report greater WMH burden in individuals from more deprived areas^13^, whereas others find no associations^16^.

A few previous studies have explored whether neighborhood deprivation relates differently to cognition and brain structure depending on individual-level factors, including race, ethnicity, sex, and genetic susceptibility. For instance, neighborhood deprivation was associated with lower cognitive performance among Black and Mexican American older adults relative to White populations^17,18^ and increased subjective cognitive impairment in a preliminary study conducted among Black women only^19^. Conversely, some studies reported better cognitive outcomes among African Americans from more disadvantaged communities, potentially reflecting selective migration processes^20^. Additionally, neighborhood socioeconomic disadvantage has been implicated racial and ethnic disparities in brain structure, with African American individuals exhibiting greater atrophy compared to White participants^21^. Although less studied, sex and genetic vulnerability may also interact with neighborhood deprivation. Preliminary data have shown increased cortical thinning and worse longitudinal cognition among men only^22^, which contrasts with studies showing that females from more deprived areas are at higher risk of developing dementia^7^. Moreover, APOE ε4 carriers living in more deprived neighborhoods showed worse cognitive function compared to non-carriers^23^. Importantly, to the best of our knowledge, no study has reported interactions between neighborhood deprivation and individual-level factors in relation to AD pathologies.

This exploratory study aims to systematically investigate the intersection between ADI and social and biological factors on a comprehensive set of age- and AD-related outcomes in cognitively unimpaired older adults. We specifically aim to examine how neighborhood disadvantage relates to cognition, amyloid and tau burden, gray matter volume, and WMH across race and ethnicity, sex and APOE ε4 groups, evaluating the effect of individual-level education. Based on previous literature, we hypothesize that the relationship between ADI and AD-related outcomes will be moderated by race and ethnicity—conceptualized as a social construct reflecting differential exposure to structural and environmental inequities—and biological factors such as sex and APOE ε4 status. In particular, we hypothesize that “Black/African American/African” and “Hispanic/Latinx/Spanish” individuals, females and APOE ε4 carriers showing stronger associations between neighborhood disadvantage and AD pathology, brain structure, and cognition.

## 2 | Methods

### Cohort and participants

The U.S. Study to Protect Brain Health Through Lifestyle Intervention to Reduce Risk (U.S. POINTER) is a multi-site, randomized controlled trial designed to assess whether a multi-domain lifestyle intervention can protect cognitive function in older adults at risk for dementia. Participant recruitment and baseline assessments were conducted between 2019 and 2022, and the trial was designed as a 2-year longitudinal study. The trial is conducted across five clinical sites in the U.S.: Chicagoland, Houston, New England/Rhode Island, North Carolina, and Northern California. The study is centrally coordinated by Wake Forest University School of Medicine, with oversight from a single Institutional Review Board (sIRB) and a Data and Safety Monitoring Board (DSMB). The trial is registered at ClinicalTrials.gov (NCT03688126)^24^.

Approximately 2,000 cognitively unimpaired older adults, aged 60 to 79 years, were enrolled based on risk factors for cognitive decline, including family history of dementia, cardiovascular risk factors, and low physical activity levels. Recruitment strategies emphasized diversity and representation, incorporating targeted outreach to historically underrepresented communities.

For this study, we used baseline data from a total of 1880 participants who had available data on demographic variables (age, sex, APOE ε4 status, and education), Area Deprivation Index, and at least one Alzheimer’s disease–related outcome: cognition (n=1880), amyloid PET (n=868), WMH (n=825), tau PET (n=845), or gray matter volume (n=836). More information about the inclusion process can be found in *Supplementary Figure 1*.

### Imaging data

#### Gray matter volume

Structural MRI data were collected using one of five Siemens 3T scanners (Tim Trio, Magnetom, Verio, Skyra, or Prisma Fit). A high-resolution 3D MPRAGE sequence was acquired for each participant using imaging parameters harmonized with the ADNI-3 protocol (repetition time = 2300 ms, minimum full echo time, voxel size = 1 mm isotropic, field of view = 256 mm). Structural MRIs were processed using FreeSurfer v7.1.1, which performs automated brain segmentation, anatomical labelling, and statistical volumetric analysis based on T1-weighted MRI scans. The Desikan-Killiany atlas was used for cortical parcellation, and a meta-ROI (composite region) was created by combining entorhinal, fusiform, parahippocampal, mid-temporal, and inferior temporal regions^25^.

#### Amyloid burden (SUVRs)

Amyloid burden was assessed using [18F]Florbetaben (FBB) PET scans; participants were injected with approximately 8 mCi of tracer and scanned 90–110 minutes post-injection (4 × 5-min frames; 20-min acquisition). Image processing followed the pipeline described by Royse et al. (2021)^26^. Each PET scan was co-registered to a native-space structural MRI, acquired closest in time to the PET scan. Structural MRIs were segmented and parcellated using FreeSurfer v7.1.1, defining a cortical summary region that includes the frontal, anterior/posterior cingulate, lateral parietal, and lateral temporal regions. Standardized uptake value ratios (SUVRs) were calculated by normalizing each FBB scan to the whole cerebellum. Results were reported in SUVRs and CL.

#### Tau burden (SUVRs)

Tau burden was measured using [18F]MK-6240 PET scans; participants were injected with approximately 5 mCi of tracer and scanned 90–110 minutes post-injection (4 × 5-min frames). Image processing followed the pipeline described by Harrison et al. (2023)^27^. Each PET scan was co-registered to a native-space structural MRI, acquired closest in time to the PET scan. To minimize the influence of off-target binding in the dorsal cerebellum, an inferior cerebellar gray matter reference region was defined using the SUIT template and reverse-normalized to native space, as described in Baker et al. (2017)^28^. SUVRs were calculated by intensity normalizing to the inferior cerebellar gray matter and extracting mean uptake values for the temporal meta-ROI (Jack et al., 2017)^29^.

PET scanners used to acquire amyloid and tau data included GE Discovery MI or ST, Siemens Biograph mCT 40 or Vision, and Phillips Gemini TF *White matter hyperintensities volume.* Segmentation was done using a pipeline that integrates T1-weighted and FLAIR MRI sequences. The segmentation was performed in minimal deformation template space using a Bayesian algorithm that identified hyperintense regions as voxels with intensity values >2.5 standard deviations above the FLAIR intensity mode, combined with spatial priors reflecting typical WMH distributions. The identified regions were then inverse-transformed to native T1 space and underwent manual quality control^30^.

### Cognitive evaluation

Preclinical Alzheimer’s Cognitive Composite (PACC) score was computed as described in previous literature^31^. In brief, this score is calculated as the sum of z-scores from four cognitive measures: MMSE (0–30), WMS-R Logical Memory Delayed Recall (0–25), Digit-Symbol Coding Test (0–93), and FCSRT96 Free + Total Recall (0–96).

### Demographic and genetic data

Demographic information, including sex, race and ethnicity, education, was collected through self-report questionnaires at enrolment. Participants selected their sex (female or male) and race and ethnicity from the following options: “Hispanic/Latinx/Spanish”, “Asian/Asian American”, “Black/African American/African”, Middle East/North African, American Indian/Alaska Native, Native Hawaiian/Other Pacific Islander, “White/European American”, or “Other”. Due to the low sample size of Middle East/North African, American Indian/Alaska Native and Native Hawaiian/Other Pacific Islander participants, these categories were combined into “Other”.

Educational attainment was self-reported via questionnaire as the highest degree completed and subsequently converted into total years of education for analysis.

APOE ε4 genotyping was performed by the Biomarker Core of the Alzheimer’s Therapeutic Research Institute (ATRI) at the University of Southern California from blood specimens collected for banking. DNA was extracted from banked blood specimens and genotyped following established ATRI procedures. Subjects were classified as carriers (at least one ε4 allele) or non-carriers (no ε4 alleles).

### Area deprivation Index

The Area deprivation index is a composite measure of neighborhood-level socioeconomic disadvantage originally developed by the U.S. Health Resources and Services Administration and subsequently refined, validated, and adapted to the Census Block Group level by the University of Wisconsin–Madison. The ADI incorporates multiple indicators including income, education, employment, and housing quality, derived from U.S. Census American Community Survey data. Neighborhoods are defined at the Census Block Group level, allowing for geographic characterization of socioeconomic context.

In this study, ADI values were obtained from the Neighborhood Atlas 2020 Zip+4 ADI Files (version 3.2), which link Census Block Group–level ADI scores to 9-digit ZIP code centroids. Participants’ residential 9-digit ZIP codes at baseline enrollment in the U.S. POINTER study were geocoded and merged with the ADI database to assign neighborhood deprivation scores. Each participant was assigned percentile ADI ranking (range 1–100) at the nation level, with higher values indicating greater neighborhood disadvantage/deprivation (100 = most disadvantaged/deprived neighborhoods).

For statistical analysis, ADI was examined both as a continuous and a categorical variable. Consistent with previous work^32^, the categorical ADI variable was divided into three groups: Low ADI (least disadvantaged, 0-30), Mid ADI (30-70), and High ADI (most disadvantaged, 70-100).

More information about the reasoning behind using ADI as a continuous and categorical variable can be found in the statistical analysis section.

## Statistical analysis

Demographic data were summarized as means (and standard deviations) for normally distributed continuous variables, medians (interquartile ranges) for skewed variables (just white matter hyperintensities), and frequencies (with percentages) for categorical variables. To characterize the cohort and aid interpretation of results, we conducted analyses to describe (1) the distribution of race/ethnicity and sex with ADI (Low, Moderate, High) using multinomial logistic regression and (2) variation in AD-related outcomes across ADI levels within each specific racial/ethnic and sex strata (using one-way ANOVA or Fisher’s exact tests).

To assess the main objective of the study—whether the relationship between ADI and AD-related outcomes (Amyloid SUVR, Tau SUVR, gray matter volume measured in a meta-ROI, WMH burden and cognition measured using PACC) varied by sex, APOE ε4 status, or race and ethnicity—we fitted multivariable linear regression models including interaction terms. Analyses were conducted (1) modelling ADI as continuous and categorical variable and (2) in two steps, first without and second with education in the model (*AD-related outcome* ∼ *ADI×sex + ADI×APOE* ε*4 status + ADI×race and ethnicity + age (+education)*). The aim of these analyse were (1) to clarify ADI associations—continuous ADI reflects a linear link with the outcome, while categorical ADI indicates differential effects across ADI levels, and to account for potential sample composition effects (e.g., under- or over-representation of a racial/ethnic group within an ADI level); (2) to report the role of education in the associations between ADI and AD related outcomes. Treating ADI as continuous variable, maximizes power and assumes a linear relationship, but it may mask group-specific differences if biomarker values differ across individual-level characteristics (see *Supplementary Tables 1, 2 and 3*). Treating ADI as categorical variable can reveal inflections and highlight localized interactions within ADI levels that continuous models may average out, but it is more susceptible to artifacts from small cell sizes and unequal group distributions. Given the exploratory nature of the analyses, stratified results by ADI category (low, moderate and high deprivation) or by the individual factor (race/ethnicity, sex and APOE ε4), using type III ANOVA, were carried out when the interaction term reached significant (p<0.05) or showed a trend (p<0.1) with the aim of facilitating interpretation of potential heterogeneity in associations across subgroups. In analyses including race and ethnicity, “White/European American” participants were used as the reference group, given that they represented the largest subgroup in the sample.

Prior to modelling, outliers were identified through visual inspection and confirmed using the interquartile range (IQR) method, with values falling below Q1 –1.5×IQR or above Q3 + 1.5×IQR excluded. This procedure aimed to reduce the influence of extreme values on model estimates. We excluded 18 outliers for white matter hyperintensities and 29 for tau SUVR; no outliers were observed for the other variables.

All analyses were conducted using R (version 4.2.2). Statistical significance was defined as a two-sided p-value < 0.05. Given the exploratory nature of the study results are interpreted in a hypothesis-generating manner. We focus on estimation and consistency with prior literature rather than strict dichotomous significance testing. Findings with p-values between 0.05 and 0.10 are reported as suggestive but not formally significant and are considered indicative of trends (borderline significant). No correction for multiple comparisons was applied.

## 3 | Results

The descriptive statistics are presented in *Table 1*. When examining the full cohort (participants with available PACC scores, N=1880) and imaging cohort (N=868), there was a higher proportion of females and “White/European American” participants within both cohorts, further the proportion of APOE ε4 carriers was 31.2% and 30.1% respectively. When comparing the full cohort and imaging cohort, the only significant difference observed between was in the sex distribution, with a lower proportion of females in the imaging cohort (χ*² = 16.05, df = 1, p < 0.001*).

**Table 1.** Demographic characteristics, socioeconomic context, cognitive performance, and Alzheimer’s disease–related biomarkers of participants in the cognition (N = 1,880) and imaging (N = 868) cohorts. Values are presented as mean (standard deviation) for continuous variables and number (percentage) for categorical variables. ADI = Area Deprivation Index; APOE = apolipoprotein E; PACC = Preclinical Alzheimer Cognitive Composite; WMH = white matter hyperintensities; SUVR = standardized uptake value ratio; meta-ROI = meta–region of interest.

|  | Cognition |  | Imaging |  |
| --- | --- | --- | --- | --- |
|  | *N=1880* | N | *N=868* | N |
| Age | 68.3 (5.16) | 1880 | 68.5 (5.24) | 868 |
| Sex: |  | 1880 |  | 868 |
| Male | 603 (32.1%) |  | 347 (40.0%) |  |
| Female | 1277 (67.9%) |  | 521 (60.0%) |  |
| Race and Ethnicity: |  | 1880 |  | 868 |
| White/European American | 1336 (71.1%) |  | 598 (68.9%) |  |
| Black/African American/African | 276 (14.7%) |  | 119 (13.7%) |  |
| Asian/Asian American | 48 (2.55%) |  | 28 (3.23%) |  |
| Hispanic/Latinx/Spanish | 106 (5.64%) |  | 64 (7.37%) |  |
| Other | 114 (6.06%) |  | 59 (6.80%) |  |
| APOE ε4 status: |  | 1880 |  | 868 |
| Carrier | 586 (31.2%) |  | 261 (30.1%) |  |
| Non-Carrier | 1294 (68.8%) |  | 607 (69.9%) |  |
| ADI level: |  | 1880 |  | 868 |
| Low | 924 (49.1%) |  | 463 (53.3%) |  |
| Moderate | 761 (40.5%) |  | 329 (37.9%) |  |
| High | 195 (10.4%) |  | 76 (8.76%) |  |
| Education (years) | 16.4 (2.14) | 1880 | 16.3 (2.12) | 868 |
| PACC | 0.00 (1.01) | 1880 | -0.08 (1.03) | 868 |
| WMH | - | - | 0.80 [0.43-1.36] | 825 |
|  | *N=1880* | N | *N=868* | N |
| Amyloid SUVR | - | - | 1.07 (0.16) | 868 |
| Tau SUVR | - | - | 1.06 (0.16) | 845 |
| Gray matter volume (meta-ROI) | - | - | 13149 (1536) | 836 |

### Sociodemographic characteristics

Sociodemographic differences in the relative probability of residing in more versus less deprived neighborhoods levels are provided in *Table 2*. In brief, participants residing in more deprived areas were more likely to be females and “Black/African American/African”. Participants who self-identified as “White/European American” or “Asian/Asian American” were more likely to reside in less deprived areas compared to other racial and ethnic groups.

**Table 2.** Relative Risk Ratio by race and sex for residing Moderate and High ADI areas compared to Low ADI areas by race and ethnicity and sex. ADI: Area Deprivation Index, RRR: Relative Risk Ratio, CI: Confidence Interval, Std.: Standard.

| Race and Ethnicity | ADI Level<br>(vs Low) | RRR | CI<br>Low | CI<br>High | Estimate | Std.<br>Error | z-value | p-value |
| --- | --- | --- | --- | --- | --- | --- | --- | --- |
| Black/African American/African | Moderate | 3.630 | 2.580 | 5.107 | 1.289 | 0.174 | 7.400 | 0.000 |
| Black/African American/African | High | 16.580 | 11.114 | 24.734 | 2.808 | 0.204 | 13.761 | 0.000 |
| Asian/Asian American | Moderate | 0.319 | 0.158 | 0.646 | -1.142 | 0.360 | -3.173 | 0.002 |
| Asian/Asian American | High | 0.124 | 0.017 | 0.906 | -2.091 | 1.017 | -2.057 | 0.040 |
| Hispanic/Latinx/Spanish | Moderate | 1.407 | 0.927 | 2.135 | 0.341 | 0.213 | 1.605 | 0.109 |
| Hispanic/Latinx/Spanish | High | 1.314 | 0.681 | 2.536 | 0.273 | 0.335 | 0.815 | 0.415 |
| Other or multiple races | Moderate | 1.020 | 0.684 | 1.522 | 0.020 | 0.204 | 0.098 | 0.922 |
| Other or multiple races | High | 0.927 | 0.476 | 1.803 | -0.076 | 0.340 | -0.224 | 0.823 |
| White/European American | Moderate | 0.558 | 0.447 | 0.696 | -0.584 | 0.113 | -5.160 | 0.000 |
| White/European American | High | 0.162 | 0.117 | 0.225 | -1.820 | 0.168 | -10.824 | 0.000 |
| Sex | ADI Level<br>(vs Low) | RRR | CI<br>Low | CI<br>High | Estimate | Std.<br>Error | z-value | p-value |
| Female | Moderate | 1.620 | 1.318 | 1.991 | 0.482 | 0.105 | 4.582 | 0.000 |
| Female | High | 3.091 | 2.078 | 4.598 | 1.128 | 0.203 | 5.570 | 0.000 |
| Male | Moderate | 0.617 | 0.502 | 0.759 | -0.482 | 0.105 | -4.580 | 0.000 |
| Male | High | 0.324 | 0.218 | 0.482 | -1.128 | 0.203 | -5.567 | 0.000 |

Demographics by ADI level and race and ethnicity, sex, and APOE ε4 status are provided in *Supplementary Tables 1, 2,* and *3*. In brief, “Black/African American/African”, “Hispanic/Latinx/Spanish”, and “White/European American” participants residing in areas with higher ADI showed fewer years of education (all p < 0.001). Higher ADI was associated with lower education among women and men, APOE ε4 carriers and non-carriers (p < 0.001). No significant differences in age were found across ADI for any sub-group.

### Interaction between ADI, Race and Ethnicity, Sex, and APOE ε4 on cognitive performance

Only race and ethnicity demonstrated a significant interaction with ADI on cognition (*Figure 1 A and B*), whereas no significant interactions were observed between ADI and APOE ε4 or sex (*Figure 1 C - F*).

**Figure 1.**
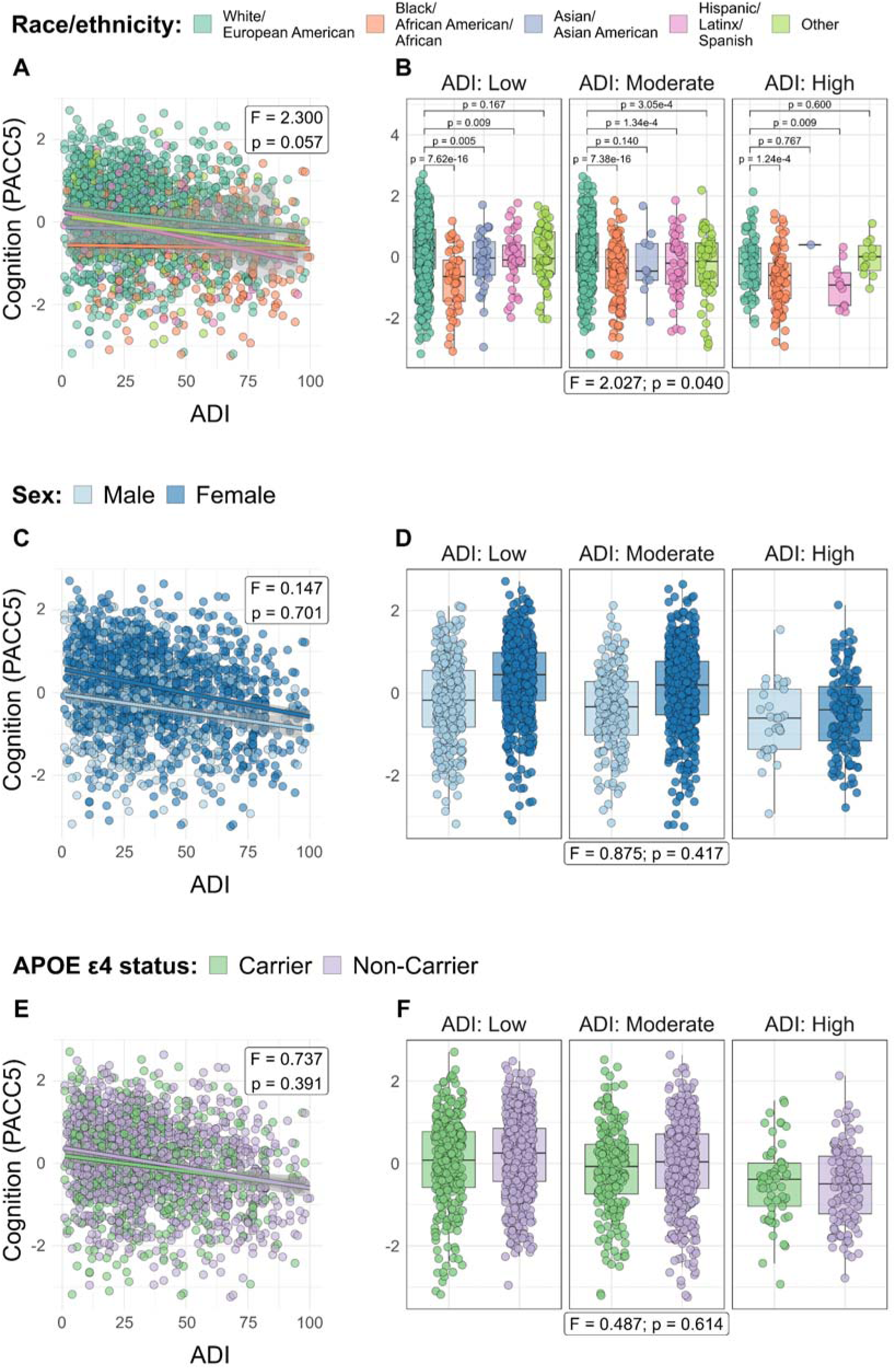
Interaction between Area Deprivation Index and race and ethnicity, sex and APOE ε4 status on cognition (PACC5). **A.** Scatter plot with fitted linear regression lines showing the relationship between continuous ADI and PACC5 scores across racial and ethnic groups. Number of subjects included in each group: White/European American: 1336; Black/African American/African: 276; Asian/Asian American: 48; Hispanic/Latinx/Spanish: 106; Other: 114. **B.** Box plots showing cognitive performance across racial and ethnic groups stratified by Area Deprivation Index level **C.** Scatter plot with fitted linear regression lines showing the relationship between continuous ADI and PACC5 scores across sex groups. Number of subjects included in each group: Males: 603; Females: 1277. **D.** Box plots showing cognitive performance across sex groups stratified by Area Deprivation Index level **E.** Scatter plot with fitted linear regression lines showing the relationship between continuous ADI and PACC5 scores across APOE ε4 status groups. Number of subjects included in each group: Carriers: 586; Non-Carriers: 1294. **F.** Box plots showing cognitive performance across APOE ε4 status groups stratified by Area Deprivation Index level Notes: In scatter plots, the shaded areas represent the 95% confidence intervals. In boxplots, ADI was divided in three levels: Low, Moderate, and High. Boxes represent the interquartile range, horizontal lines indicate medians, and whiskers denote 1.5× the interquartile range. P-values between groups are indicated for the significant interactions. Number of subjects included in each ADI level: Low: 924; Moderate: 761; High: 195. P-values for comparisons in boxplots were only shown when the overall interaction was significant or a trend (p<0.05 or p<0.1 respectively).

Specifically, a trend towards an interaction was observed between continuous ADI and race and ethnicity on cognition (*Figure 1A*), (*Supplementary Table 4*). When studying the association between ADI and cognition within each racial and ethnic group we found that among “White/European American”, “Hispanic/Latinx/Spanish”, and “Other” participants, higher continuous ADI was significantly associated with lower PACC scores, meanwhile the remaining racial and ethnic groups did not show a significant association (*Supplementary Table 5*). Further, treating ADI as categorial, a significant interaction was observed with race and ethnicity on PACC (*Supplementary Table 6*) such that the cognitive performance gap between “White/European American” and “Black/African American/African” participants was larger in low-ADI areas and narrowed at higher ADI levels, reflecting stable lower PACC scores across ADI levels among “Black/African American/African” participants and lower cognitive performance with higher ADI among “White/European American” participants. In contrast, the cognitive gap between “White/European American” and “Hispanic/Latinx/Spanish” participants widened at higher ADI levels, as “Hispanic/Latinx/Spanish” participants living in more deprived areas exhibited lower cognitive scores compared with “White/European American” participants from the same ADI level. Moreover, “Asian/Asian American” in low-ADI areas and “Other” participants in moderately deprived areas also showed lower scores compared to “White/European American” participants (*Figure 1B*) (*Supplementary Table 7*).

Notably, Accounting for education reduced the significance of the overall interaction between ADI and race and ethnicity on cognition to non-significant for ADI continuous and borderline for ADI categorical, indicating that educational attainment may partly explain the observed racial and ethnic heterogeneity in the association between neighborhood disadvantage and cognitive performance (*Supplementary Tables 8* and *9*). At the group level, the relationship between ADI continuous and cognition became non-significant for “Hispanic/Latinx/Spanish” and remained unchanged for “White/European American” and “Other” participants. (*Supplementary Table 10).* Across ADI levels, adjusting by education influenced the results in highly deprived areas (attenuating the difference between “White/European American” and “Hispanic/Latinx/Spanish” to non-significant) while the overall pattern remained similar in less and moderately deprived areas (*Supplementary Table 11*).

### Interaction between ADI and Race and Ethnicity, Sex, and APOE ε4 on AD pathology

No significant interactions were observed with race and ethnicity, sex and APOE ε4 and continuous ADI on AD pathology. Regarding ADI categorical, APOE ε4 status and sex showed significant differential associations with AD pathologies across ADI levels (amyloid results shown in *Figure 2* and tau results in *Figure 3*).

**Figure 2.**
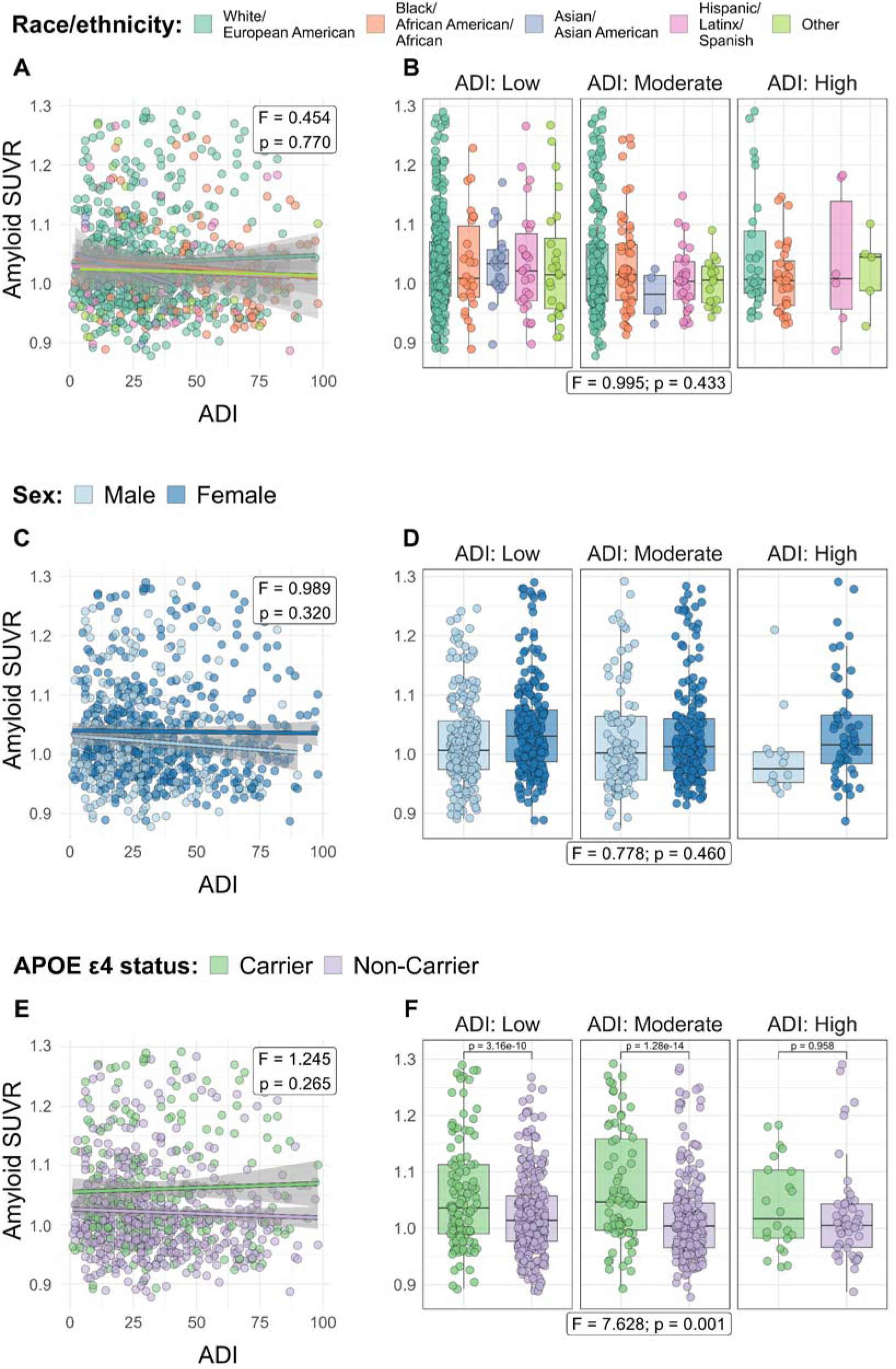
Interaction between Area Deprivation Index and race and ethnicity, sex and APOE ε4 on amyloid SUVR. **A.** Scatter plot with fitted linear regression lines showing the relationship between continuous ADI and amyloid SUVR across racial and ethnic groups. Number of subjects included in each group: White/European American: 598; Black/African American/African: 119; Asian/Asian American: 28; Hispanic/Latinx/Spanish: 64; Other: 59. **B.** Box plots showing amyloid SUVR across racial and ethnic groups stratified by Area Deprivation Index level **C.** Scatter plot with fitted linear regression lines showing the relationship between continuous ADI and amyloid SUVR across sex groups. Number of subjects included in each group: Males: 347; Females: 521. **D.** Box plots showing amyloid SUVR across sex groups stratified by Area Deprivation Index level **E.** Scatter plot with fitted linear regression lines showing the relationship between continuous ADI and amyloid SUVR across APOE ε4 status groups. Number of subjects included in each group: Carriers: 261; Non-Carriers: 607. **F.** Box plots showing amyloid SUVR across APOE ε4 status groups stratified by Area Deprivation Index level Notes: In scatter plots, the shaded areas represent the 95% confidence intervals. In boxplots, ADI was divided in three levels: Low, Moderate, and High. Boxes represent the interquartile range, horizontal lines indicate medians, and whiskers denote 1.5× the interquartile range. P-values between groups are indicated for the significant interactions. Number of subjects included in each ADI level: Low: 463; Moderate: 329; High: 76.

**Figure 3.**
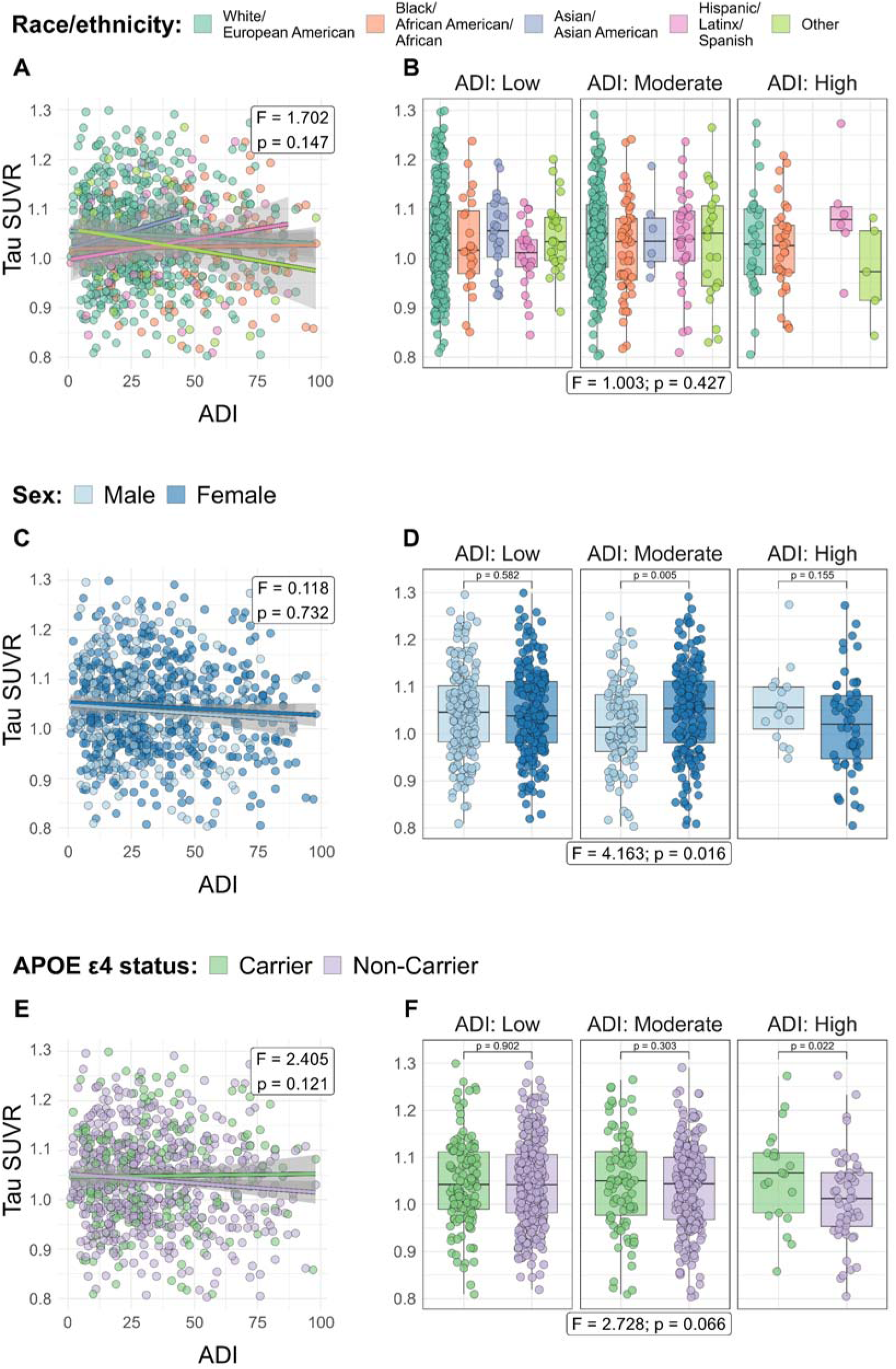
Interaction between Area Deprivation Index and race and ethnicity, sex and APOE ε4 on tau SUVR. **A.** Scatter plot with fitted linear regression lines showing the relationship between continuous ADI and tau SUVR across racial and ethnic groups. Number of subjects included in each group: White/European American: 561; Black/African American/African: 112; Asian/Asian American: 27; Hispanic/Latinx/Spanish: 62; Other: 54. **B.** Box plots showing tau SUVR across racial and ethnic groups stratified by Area Deprivation Index level **C.** Scatter plot with fitted linear regression lines showing the relationship between continuous ADI and tau SUVR across sex groups. Number of subjects included in each group: Males: 327; Females: 489. **D.** Box plots showing tau SUVR across sex groups stratified by Area Deprivation Index level **E.** Scatter plot with fitted linear regression lines showing the relationship between continuous ADI and tau SUVR across APOE ε4 status groups. Number of subjects included in each group: Carriers: 240; Non-Carriers: 576. **F.** Box plots showing tau SUVR across APOE ε4 status groups stratified by Area Deprivation Index level Notes: In scatter plots, the shaded areas represent the 95% confidence intervals. In boxplots, ADI was divided in three levels: Low, Moderate, and High. Boxes represent the interquartile range, horizontal lines indicate medians, and whiskers denote 1.5× the interquartile range. P-values between groups are indicated for the significant interactions. Number of subjects included in each ADI level: Low: 431; Moderate: 312; High: 73.

Thus, while no significant interaction was found between continuous ADI and APOE ε4 status on amyloid (*Supplementary Table 12*), a significant interaction was observed between ADI categorical and APOE ε4 status on amyloid (*Supplementary Table 13*) (*Figure 2E* and *Figure 2F*, respectively). Among participants living in low and moderately deprived areas amyloid burden differed by APOE ε4 status, with higher amyloid burden observed in APOE ε4 carriers compared to non-carriers, whereas no such difference was found among those living in highly deprived areas (*Supplementary Table 14*). No significant interaction between ADI categorical and sex or race and ethnicity on amyloid was found.

Including education in the models did not affect these results (*Supplementary Tables 15* and *16*). Importantly, these models were also ran using centiloids as outcome instead of SUVRs and the results were the same (*Supplementary Tables 17-20*).

Continuous ADI did not interact with race and ethnicity, sex or APOE ε4 status on tau (*Supplementary Table 21*; *Figures 3A, 3C* and *3E*, respectively). However, we observed a trend toward an interaction between categorical ADI and APOE ε4 status on tau and a significant interaction between ADI and sex (*Supplementary Table 22; Figures 3F* and *3D* respectively). The interactions indicated that among participants living in highly deprived areas, APOE ε4 carriers exhibited significantly higher tau compared to non-carriers (*Supplementary Table 23*) and that female from moderately deprived areas showed higher tau levels compared to males (*Supplementary Table 23*).

Both interactions remained unchanged when education was controlled for (*Supplementary Tables 24* and *25*).

### Interaction between ADI and Race and Ethnicity, Sex, and APOE ε4 on white matter hyperintensities and gray matter volume

Continuous ADI interacted with APOE ε4 status and race and ethnicity on WMH. Differential associations of APOE ε4 on WMH were also observed across ADI levels (WMH results shown in *Figure 4*) (*Supplementary Tables 26 and 27*).

**Figure 4.**
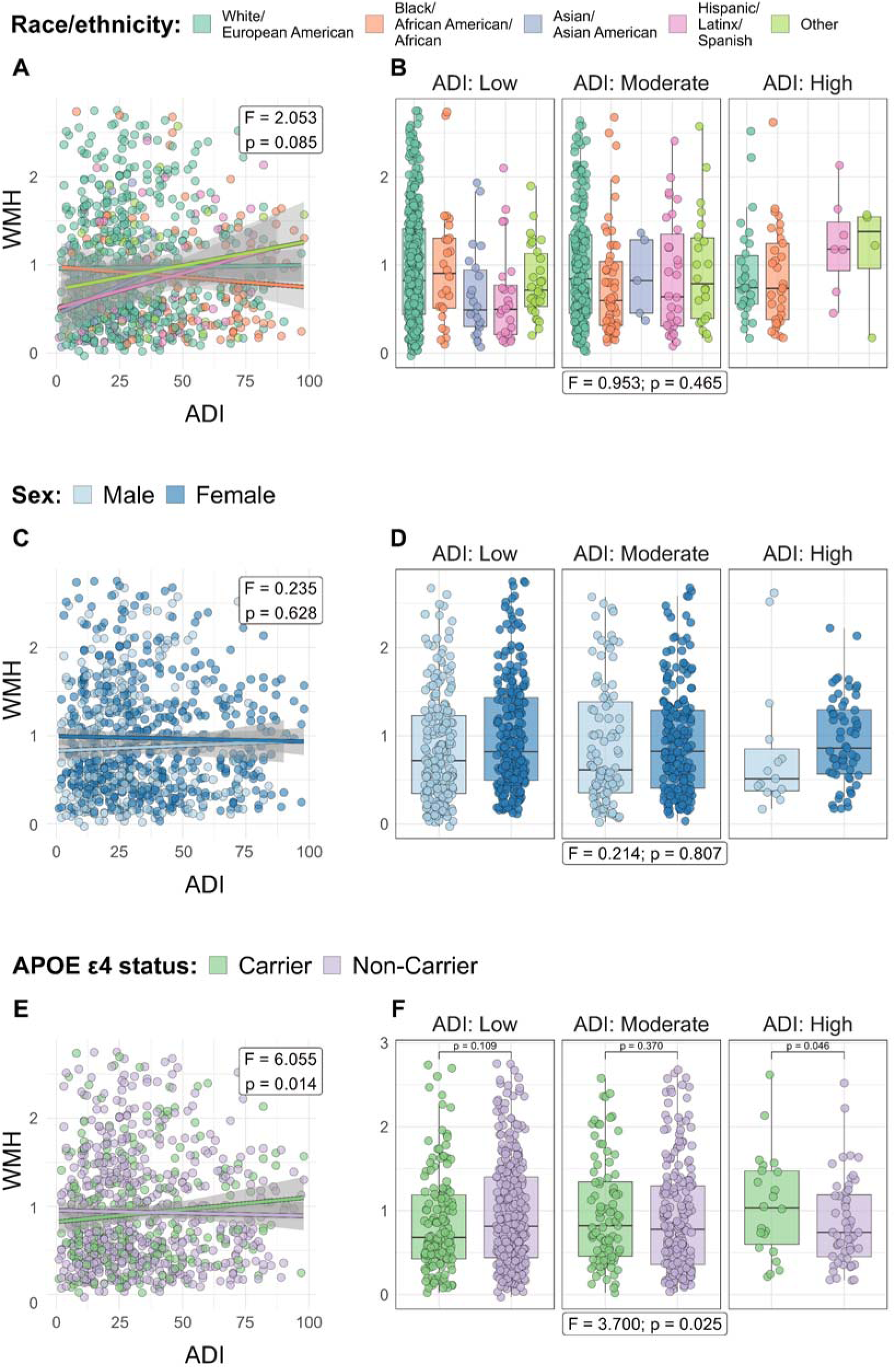
Interaction between Area Deprivation Index and race and ethnicity, sex and APOE ε4 on WMH. **A.** Scatter plot with fitted linear regression lines showing the relationship between continuous ADI and WMH across racial and ethnic groups. Number of subjects included in each group: White/European American: 574; Black/African American/African: 112; Asian/Asian American: 27; Hispanic/Latinx/Spanish: 61; Other: 56. **B.** Box plots showing WMH across racial and ethnic groups stratified by Area Deprivation Index level **C.** Scatter plot with fitted linear regression lines showing the relationship between continuous ADI and WMH across sex groups. Number of subjects included in each group: Males: 330; Females: 500. **D.** Box plots showing WMH across sex groups stratified by Area Deprivation Index level **E.** Scatter plot with fitted linear regression lines showing the relationship between continuous ADI and WMH across APOE ε4 status groups. Number of subjects included in each group: Carriers: 255; Non-Carriers: 575. **F.** Box plots showing WMH across APOE ε4 status groups stratified by Area Deprivation Index level Notes: In scatter plots, the shaded areas represent the 95% confidence intervals. In boxplots, ADI was divided in three levels: Low, Moderate, and High. Boxes represent the interquartile range, horizontal lines indicate medians, and whiskers denote 1.5× the interquartile range. P-values between groups are indicated for the significant interactions. Number of subjects included in each ADI level: Low: 450; Moderate: 306; High: 74.

Thus, a significant interaction between continuous ADI and APOE ε4 status on WMH indicated that among APOE ε4 carriers, higher ADI was significantly associated with greater WMH burden (*Supplementary Table 28; Figure 4E*). In line with this, only APOE ε4 carriers residing in highly deprived areas exhibited significantly higher levels of WMH compared to non-carriers, as observed in models using categorial ADI (*Supplementary Tables 29*; *Figure 4F*). These interactions remained significant after adjusting for education, although the ADI-WMH association within APOE ε4 carriers was attenuated to a borderline effect (*Supplementary Tables 30-33*).

Continuous ADI also showed a borderline interaction with race and ethnicity on WMH (*Figure 4A*; *Supplementary Table 26*) such that higher ADI showed a trend toward higher WMH burden among “Hispanic/Latinx/Spanish” and “Other” participants (*Supplementary Table 34*). No significant interactions were observed between ADI categorical and race and ethnicity on WMH (*Supplementary Table 27*; *Figure 4B*). After adjusting for education, the interaction between ADI continuous and APOE ε4 status on WMH remained borderline (*Supplementary Tables 30*); however, the association between ADI and WMH became significant among “Hispanic/Latinx/Spanish” (*Supplementary Table 35*).

No significant interaction between continuous or categorical ADI and sex on WMH was found (*Supplementary Table 26 and 27*).

No significant interactions were observed on GM volume when ADI was treated as either continuous or categorical (*Supplementary Tables 36* and *37*) (GM volume results shown in *Figure 5*).

**Figure 5.**
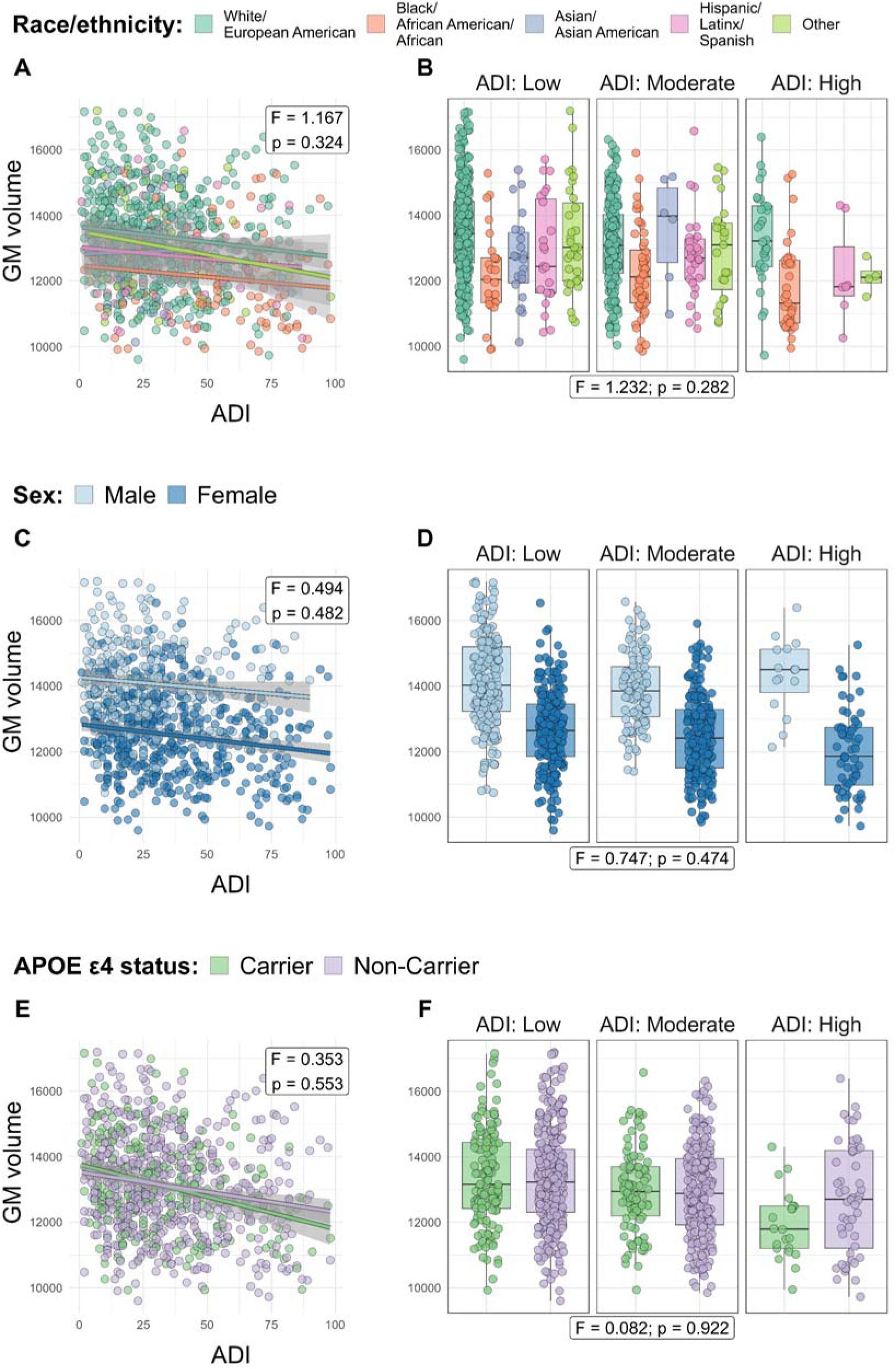
Interaction between Area Deprivation Index and race and ethnicity, sex and APOE ε4 on GM volume. **A.** Scatter plot with fitted linear regression lines showing the relationship between continuous ADI and GM volume across racial and ethnic groups. Number of subjects included in each group: White/European American: 579; Black/African American/African: 111; Asian/Asian American: 28; Hispanic/Latinx/Spanish: 60; Other: 58. **B.** Box plots showing GM volume across racial and ethnic groups stratified by Area Deprivation Index level **C.** Scatter plot with fitted linear regression lines showing the relationship between continuous ADI and GM volume across sex groups. Number of subjects included in each group: Males: 331; Females: 505. **D.** Box plots showing GM volume across sex groups stratified by Area Deprivation Index level **E.** Scatter plot with fitted linear regression lines showing the relationship between continuous ADI and GM volume across APOE ε4 status groups. Number of subjects included in each group: Carriers: 253; Non-Carriers: 583. **F.** Box plots showing GM volume across APOE ε4 status groups stratified by Area Deprivation Index level Notes: In scatter plots, the shaded areas represent the 95% confidence intervals. In boxplots, ADI was divided in three levels: Low, Moderate, and High. Boxes represent the interquartile range, horizontal lines indicate medians, and whiskers denote 1.5× the interquartile range. P-values between groups are indicated for the significant interactions. Number of subjects included in each ADI level: Low: 448; Moderate: 314; High: 74.

## 4 | Discussion

In a predominantly unimpaired clinical trial cohort enriched for geographic and ethnoracial diversity, we found preliminary, moderate evidence that the interplay of individual-level social and biological factors with neighborhood deprivation varied across AD-related outcomes. Specifically, (i) ADI interacted with race and ethnicity on cognition: “White/European American”, “Hispanic/Latinx/Spanish”, and “Other” showed lower cognitive scores with higher ADI, however, across ADI levels, “Black/African American/African” and “Hispanic/Latinx/Spanish” participants showed lower cognitive scores than “White/European American”, with education attenuating these associations; (ii) ADI level-specific associations were observed for amyloid and tau, with females from moderately deprived areas presenting higher tau burden, while APOE ε4 carriers exhibited differing patterns –higher amyloid burden in less deprived areas and a trend towards higher tau in more deprived areas; (iii) race and ethnicity and APOE ε4 status interacted with ADI on WMH, with higher ADI associated with greater WMH burden among “Hispanic/Latinx/Spanish” participants and APOE ε4 carriers.

Taken together, our results provide preliminary evidence that neighborhood deprivation, in combination with social and biological factors, may increase vulnerability to specific AD-related outcomes. Across the two analytical approaches, the most consistent findings were: race and ethnicity influencing ADI-cognition and WMH associations. Less consistent findings pointed to female sex affecting tau, and APOE ε4 carriership modulating the ADI-WMH association. Notably, only the APOE ε4 moderation of ADI-amyloid survived multiple comparison correction (*Supplementary Table 38*). Given the exploratory nature of this study, we interpret results cautiously. These patterns likely reflect systemic health disparities and sample composition or recruitment, highlighting that observed effects are shaped by both structural inequities as well as study design.

Race and ethnicity influenced the relationship between ADI and cognition. Within racial and ethnic groups, higher ADI was associated with lower cognitive performance among “White/European American”, “Hispanic/Latinx/Spanish” but not among “Black/African American/African” participants. The absence of an ADI–cognition association in “Black/African American/African” participants may suggest that factors beyond ADI are influencing cognition in this group. Indeed, when examining cognition within each ADI level, both “Hispanic/Latinx/Spanish” and “Black/African American/African” participants showed significantly lower cognitive performance compared to “White/European American”. This disparity may reflect the enduring effect of structural racism and lifetime exposure to disadvantage –factors not fully captured by ADI– indicating that inequalities persist even in low-ADI neighborhoods. This interpretation aligns with studies reporting lower cognitive scores among Hispanic^33^ and African American^34,35^ individuals compared to White populations, attributed to disparities in educational opportunities and structural inequities.

Previous studies have shown lower cognitive scores among “Hispanic/Latinx/Spanish” and “Black/African American/African” older adults in more disadvantaged areas^17,18^, with one study observing better performance among African American individuals from disadvantaged environments^19^, likely reflecting historical migration patterns. The differing results may be explained by sample composition or recruitment bias, with underrepresentation of “Black/African American/African” participants in low-ADI areas and disproportionate concentration in high ADI neighborhoods. This pattern may have limited our ability to detect an association between ADI and cognition within this group and reduced statistical power. Further, the underrepresentation of higher-functioning “Black/African American/African” participants in low-ADI neighborhoods or “White/European American” participants in high-ADI neighborhoods could make the ADI–cognition association appear steeper among “White/European American” participants. These factors should be considered when interpreting the results.

Accounting for individual-level education attenuated the interaction between ADI and race on cognition, diminishing the effect of ADI on cognition among “Hispanic/Latinx/Spanish” and “Other” participants, and reducing the differences in cognitive performance between “White/European American” and “Hispanic/Latinx/Spanish” participants from high ADI areas. This finding aligns with prior studies showing that higher educational attainment can buffer against the negative effects of neighborhood deprivation on cognition^36,37^.

APOE ε4 status influenced AD pathology and WMH only within specific ADI strata, suggesting a gene-environment interaction in which the effect of genetic susceptibility varies according to neighborhood context. Specifically, we observed higher WMH and tau burden among carriers from highly deprived. Unexpectedly, we also found increased amyloid burden among carriers from less deprived areas. The APOE ε4 by ADI interaction on tau and amyloid emerged only when ADI was modelled categorically, suggesting that AD-pathology vulnerability in carriers may be nonlinear, becoming evident at specific ADI levels. In contrast, the APOE ε4 by ADI interaction on WMH was evident in both categorical and continuous analyses, indicating a more linear relationship.

Higher tau and WMH in carriers from highly deprived areas, together with the absence of increased amyloid in carriers from more deprived areas, may reflect the interplay of multiple, potentially competing, pathways. Elevated vascular and metabolic risks in high-ADI neighborhoods^38,39^ in APOE ε4 carriers are consistent with the greater WMH burden observed, suggesting that socioeconomic disadvantage shapes neurodegenerative processes through both vascular and tau-related mechanisms rather than through amyloid alone. The absence of increased amyloid in APOE ε4 carriers from more deprived backgrounds might be associated with cognitive resilience against amyloid deposition^40,41^. Carriers from less deprived neighborhoods may present higher amyloid without developing clinical dementia, coping with pathological burden^42,43^. Post-hoc analysis, including PACC scores in this model, supported this interpretation that carriers in low-ADI areas showed higher amyloid independent of their cognitive level (data not shown).

Among APOE ε4 carriers, education attenuated the association between ADI and WMH, suggesting that individual-level educational attainment may buffer the effect of ADI on WMH in APOE ε4 carriers. This aligns with other studies suggesting the attenuating role of education on brain outcomes in APOE ε4 carriers^44^. Previous studies have shown that APOE ε4 carriers exhibit greater tau pathology^45^ and higher neighborhood deprivation is associated with increased tau burden^14^. Our results integrate these findings by suggesting a gene-environment interaction. Findings from previous studies regarding WMH and deprivation are mixed; some studies reported stronger associations^13^, while others found no significant relationship^16^. Our results suggest that the relationship between ADI and WMH may be more pronounced in APOE ε4 carriers individuals, underscoring the need to study the effect of ADI in this population.

Taken together, results related to APOE ε4 status suggest that neighborhood deprivation may interact differently with genetic risk across AD biomarkers: advantaged areas may confer resilience to amyloid—potentially through higher education and cognitive reserve^46,47^—whereas greater deprivation may exacerbate tau and WMH burden, possibly linked to poorer cardiovascular health, which is common among individuals from more deprived areas^38,39^.

Sex influenced the ADI–tau relationship. A significant interaction emerged only with ADI categorical, suggesting effects at specific deprivation levels rather than across the entire gradient. Females from moderately deprived areas showed higher tau burden. This effect was not observed in highly deprived areas, possibly due to the smaller number of participants in this stratum. Our findings align with previous research showing that females exhibit higher levels of tau pathology compared to males^45,48,49^ but prior studies have not examined the role of ADI in this relationship. Our study suggests that tau burden is not only influenced by sex, but neighborhood disadvantage may amplify this effect.

We also observed a borderline interaction between ADI continuous and race and ethnicity on WMH, with “Hispanic/Latinx/Spanish” individuals showing increased WMH burden with higher ADI. Given prior evidence linking Hispanic ethnicity to higher WMH burden^50^, our findings suggest that neighborhood deprivation may amplify vulnerability to WMH in this population. Adjusting for education, consistent with patterns observed across analyses, strengthened this association, indicating that individual educational attainment may partially mask neighborhood-level effects.

No significant interactions were observed with gray matter volume. This contrasts with previous research linking higher deprivation to reduced brain volume^12,13,51^ and CSF t-tau^14^. Several factors may account for these discrepancies. Prior studies have often focused on specific regions, such as the hippocampus, while we looked at an established AD-signature composite region^25^. In addition, although there is literature examining similar brain regions^51^, the effects were observed among Black participants, including individuals spanning cognitively unimpaired, MCI, and dementia stages. In contrast, our sample consisted exclusively of cognitively unimpaired older adults. Together, differences in the brain regions examined, racial sub-group effects, and clinical composition of the sample may help explain why similar interactions were not found.

This study has limitations. Given its exploratory design, we did not apply correction for multiple comparisons; therefore, results should be interpreted as preliminary and hypothesis-generating, intended to guide future research, rather than definitive conclusions. In addition, analyses were cross-sectional and based on baseline data, limiting inference on temporal relationships. Future longitudinal follow-up will be needed to assess associations with trajectories of pathology and cognitive decline. Other limitations include: reliance on residence at baseline/enrollment to assign ADI, and ADI capturing structural/contextual education rather than individual-level education quality, which may differ across racial/ethnic groups^52^. Strengths include: examination of multiple AD-related outcomes, use of validated neighborhood deprivation measure, and a well-characterized, diverse ethno-racial cohort.

In summary, this study provides preliminary evidence that neighborhood socioeconomic disadvantage may be associated with AD-related outcomes in ways that vary across individual social and biological factors. These findings extend prior work by integrating neighborhood context with cognitive and multimodal neuroimaging measures in a well-characterized cohort of cognitively unimpaired older adults. Future research should explore lifelong socioeconomic exposures and examine potential protective factors in more diverse cohorts to inform targeted interventions aimed at reducing disparities.

## Data Availability

The data used in this study were obtained from the U.S. POINTER study database and are not publicly available. Data may be made available upon reasonable request and subject to approval by the U.S. POINTER study investigators and applicable data-use agreements.

## Acknowledgements

The data used for this paper were obtained from the U.S. Study to Protect Brain Health Through Lifestyle Intervention to Reduce Risk (U.S. POINTER) database. The investigators involved in U.S. POINTER contributed to its design and implementation and/or provided data but were not involved in the analysis or writing of this paper. We gratefully acknowledge all U.S. POINTER participants, as well as the study teams and participating sites for their essential contributions. The full list of U.S. POINTER investigators can be found in the supplementary materials (*Supplementary Table 38*).

## Conflicts of Interest

SNL is a full-time employee of Perceptive, Inc.

HMS is a full-time employee of Alzheimer’s Association; her husband works at Abbott Labs in an unrelated field.

SML is on an advisory board for the NIH IPAT study and has received speaking honoraria from IMPACT-AD and J&J. She has consulted for Banner Health and received travel support from J&J, the Alzheimer’s Therapeutic Research Institute, Alzheimer’s Association, Shenzhen Bay Lab, and is on the editorial board for JAMA Neurology.

PAD, CMC, MPJ, TMH, JCM, LTVM, LDB, AB and EMAU report no conflict of interest.

## Funding Sources

NIA R01AG062689 (POINTER imaging grant)

Alzheimer’s Association: U.S. POINTER-19-611541 (main POINTER trial grant)

The research leading to these results has received funding from the Ajuntament de Barcelona (23S06157-001), Ministry of Science and Innovation (PID2019-111514RA-I00), and the Alzheimer’s Association research grants (AARG 2019-AARG-644641, AARG2019-AARG-644641-RAPID), to EMA-U. EMA-U is supported by the Spanish Ministry of Science and Innovation—State Research Agency (RYC2018-026053-I), co-funded by the European Social Fund (ESF). The sponsor had no role in the design and conduct of the study; in the collection, analysis, and interpretation of data; in the preparation of the manuscript; or in the review or approval of the manuscript.

AB acknowledges support from Instituto de Salud Carlos III and co-funded by the European Union through the Miguel Servet grant (CP20/00038) and Fondo de Investigaciones Sanitario (PI22/00307), the Alzheimer’s Association (AARG-22-923680), and the Ajuntament de Barcelona, in collaboration with Fundació La Caixa (23S06157-001).

## Notes

### Author Declarations

The Institutional Review Board of Wake Forest University School of Medicine gave ethical approval for this work. The U.S. POINTER study was conducted under oversight of a single Institutional Review Board and a Data and Safety Monitoring Board.

