## Supplemental material for "Association of neighborhood deprivation with Alzheimer’s Disease pathology, brain structure, and cognition by race and ethnicity, sex, and APOE ε4 status"

### **Supplementary Figure 1.** Graph representing final selection of subjects

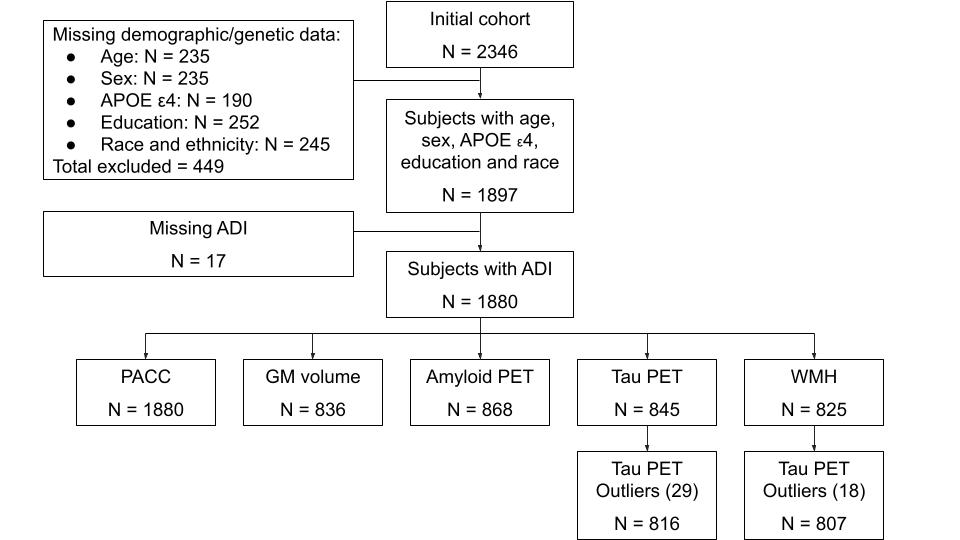

Abbreviations: APOE = Apolipoprotein E; ADI = Area Deprivation Index; PACC = Preclinical Alzheimer’s Cognitive Composite; GM = Grey Matter; PET = Positron Emission Tomography; WMH = White Matter Hyperintensities.

Note. Data was received on 27/09/2023.

### **Supplementary Table 1.** Demographic data and AD related outcomes stratified by race and ethnicity and ADI level.

|  | African American | | | | Asian | | | | Hispanic | | | | Other or multiple races | | | | White | | | |
| --- | --- | --- | --- | --- | --- | --- | --- | --- | --- | --- | --- | --- | --- | --- | --- | --- | --- | --- | --- | --- |
|  | Low | Mod | High | p | Low | Mod | High | p | Low | Mod | High | p | Low | Mod | High | p | Low | Mod | High | p |
|  | N=50 (18.1%) | N=131 (47.5%) | N=95 (34.4%) |  | N=37 (76.0%) | N=10 (20.6%) | N=1 (21.0.%) |  | N=44 (42.3%) | N=50 (48.1%) | N=12 (11.5%) |  | N=56 (48.3%) | N=47 (40.5%) | N=11 (9.5%) |  | N=737 (54.9%) | N=523 (38.9%) | N=76 (5.7%) |  |
| Age | 68.3 (4.97) | 67.3 (4.81) | 68.7 (4.85) | 0.076 | 68.1 (5.26) | 67.8 (4.61) | 63.0 (.) | 0.618 | 66.7 (4.84) | 67.1 (5.21) | 69.1 (4.78) | 0.340 | 68.2 (5.10) | 68.0 (5.88) | 68.5 (4.61) | 0.954 | 68.4 (5.26) | 68.6 (5.12) | 68.5 (5.26) | 0.923 |
| Sex: |  |  |  | 0.064 |  |  |  | 0.825 |  |  |  | 0.018 |  |  |  | 0.088 |  |  |  | 0.012 |
| Male | 12 (24.0%) | 20 (15.3%) | 9 (9.47%) |  | 15 (40.5%) | 3 (30.0%) | 0 (0.00%) |  | 20 (45.5%) | 10 (20.0%) | 2 (16.7%) |  | 23 (41.1%) | 13 (27.7%) | 1 (9.09%) |  | 287 (38.9%) | 167 (31.9%) | 21 (27.6%) |  |
| Female | 38 (76.0%) | 111 (84.7%) | 86 (90.5%) |  | 22 (59.5%) | 7 (70.0%) | 1 (100%) |  | 24 (54.5%) | 40 (80.0%) | 10 (83.3%) |  | 33 (58.9%) | 34 (72.3%) | 10 (90.9%) |  | 450 (61.1%) | 356 (68.1%) | 55 (72.4%) |  |
| APOE ε4 |  |  |  | 0.282 |  |  |  | 0.724 |  |  |  | 0.699 |  |  |  | 1.000 |  |  |  | 0.467 |
| Carrier | 23 (46.0%) | 45 (34.4%) | 32 (33.7%) |  | 8 (21.6%) | 1 (10.0%) | 0 (0.00%) |  | 11 (25.0%) | 9 (18.0%) | 2 (16.7%) |  | 18 (32.1%) | 16 (34.0%) | 4 (36.4%) |  | 235 (31.9%) | 163 (31.2%) | 19 (25.0%) |  |
| Non-Carrier | 27 (54.0%) | 86 (65.6%) | 63 (66.3%) |  | 29 (78.4%) | 9 (90.0%) | 1 (100%) |  | 33 (75.0%) | 41 (82.0%) | 10 (83.3%) |  | 38 (67.9%) | 31 (66.0%) | 7 (63.6%) |  | 502 (68.1%) | 360 (68.8%) | 57 (75.0%) |  |
| Education | 16.4 (1.98) | 16.4 (2.00) | 15.4 (2.13) | <0.001†‡ | 16.8 (1.97) | 16.8 (1.93) | 14.0 (.) | 0.375 | 16.7 (2.03) | 15.0 (2.15) | 13.3 (1.30) | <0.001 *†‡ | 16.8 (2.15) | 16.3 (2.10) | 15.6 (2.16) | 0.218 | 16.8 (1.98) | 16.2 (2.18) | 15.4 (2.24) | <0.001 *†‡ |
| PACC 5 | -0.75 (1.02) | -0.48 (0.99) | -0.72 (0.91) | 0.096 | -0.15 (0.92) | -0.24 (1.07) | 0.40 (.) | 0.814 | -0.04 (0.86) | -0.24 (1.01) | -0.92 (0.69) | 0.015  † | 0.06 (0.93) | -0.35 (1.16) | -0.04 (0.62) | 0.121 | 0.23 (0.97) | 0.10 (0.96) | -0.24 (0.90) | <0.001 *†‡ |
| WMH | 1.64 (2.01) | 1.71 (2.73) | 1.76 (2.52) | 0.986 * | 1.01 (1.33) | 1.35 (0.98) | . (.) | 0.601 | 1.07 (1.58) | 1.76 (2.13) | 2.54 (2.20) | 0.165 | 1.32 (1.14) | 2.09 (2.59) | 2.11 (1.51) | 0.315 | 1.97 (2.26) | 2.07 (2.46) | 1.84 (2.36) | 0.828 |
| Amyloid SUVR | 1.05 (0.14) | 1.04 (0.10) | 1.03 (0.12) | 0.807 | 1.03 (0.06) | 1.13 (0.24) | . (.) | 0.080 | 1.03 (0.09) | 1.04 (0.15) | 1.08 (0.16) | 0.728 | 1.12 (0.19) | 1.19 (0.32) | 1.02 (0.07) | 0.322 | 1.08 (0.15) | 1.08 (0.17) | 1.07 (0.13) | 0.840 |
| Tau SUVR | 1.06 (0.19) | 1.02 (0.09) | 1.02 (0.09) | 0.329 | 1.05 (0.08) | 1.05 (0.08) | . (.) | 0.984 | 1.01 (0.07) | 1.04 (0.10) | 1.20 (0.31) | 0.003  †‡ | 1.11 (0.26) | 1.06 (0.19) | 0.97 (0.10) | 0.389 | 1.08 (0.16) | 1.07 (0.17) | 1.06 (0.16) | 0.678 |
| GM volume (meta-ROI) | 12202 (1391) | 12158 (1425) | 11807 (1378) | 0.455 | 12691 (1346) | 13557 (1680) | . (.) | 0.196 | 13084 (1836) | 12748 (1276) | 12221 (1506) | 0.411 | 13277 (1629) | 12941 (1471) | 12124 (507) | 0.328 | 13566 (1505) | 13164 (1370) | 13332 (1464) | 0.008  * |

Note: Values are shown as mean (standard deviation) for continuous variables and number (percentage) for categorical variables. P-values for continuous variables were obtained from one-way ANOVA within each sex across ADI categories; p-values for categorical variables were obtained from Fisher’s exact test. For variables with significant overall ANOVA p-values, post-hoc pairwise comparisons with correction for multiple testing were performed (see footnotes).

Note 2: For variables with significant overall p-values, Tukey-adjusted post-hoc pairwise comparisons are indicated by symbols: * Moderate vs Low; † High vs Low; ‡ High vs Moderate (p<0.05).

Note 3: For these comparisons we used the full cohort for the variables that are present in both imaging and full cohort. For the variables only present in imaging cohort only subjects from that cohort were included.

### **Supplementary Table 2.** Demographic data and AD related outcomes stratified by sex and ADI level.

|  | **Female** | | | | **Male** | | | |
| --- | --- | --- | --- | --- | --- | --- | --- | --- |
|  | **Low** | **Moderate** | **High** | **p** | **Low** | **Moderate** | **High** | **p** |
|  | ***N=567*** | ***N=548*** | ***N=162*** |  | ***N=357*** | ***N=213*** | ***N=33*** |  |
| Age | 68.1 (5.17) | 68.0 (5.23) | 68.5 (4.91) | 0.527 | 68.6 (5.28) | 68.8 (4.85) | 69.2 (5.28) | 0.820 |
| Race: |  |  |  | <0.001 |  |  |  | <0.001 |
| White | 450 (79.4%) | 356 (65.0%) | 55 (34.0%) |  | 287 (80.4%) | 167 (78.4%) | 21 (63.6%) |  |
| African American | 38 (6.70%) | 111 (20.3%) | 86 (53.1%) |  | 12 (3.36%) | 20 (9.39%) | 9 (27.3%) |  |
| Asian | 22 (3.88%) | 7 (1.28%) | 1 (0.62%) |  | 15 (4.20%) | 3 (1.41%) | 0 (0.00%) |  |
| Hispanic | 24 (4.23%) | 40 (7.30%) | 10 (6.17%) |  | 20 (5.60%) | 10 (4.69%) | 2 (6.06%) |  |
| Other or multiple races | 33 (5.82%) | 34 (6.20%) | 10 (6.17%) |  | 23 (6.44%) | 13 (6.10%) | 1 (3.03%) |  |
| APOE ε4: |  |  |  | 0.781 |  |  |  | 0.855 |
| Carrier | 181 (31.9%) | 166 (30.3%) | 48 (29.6%) |  | 114 (31.9%) | 68 (31.9%) | 9 (27.3%) |  |
| Non-Carrier | 386 (68.1%) | 382 (69.7%) | 114 (70.4%) |  | 243 (68.1%) | 145 (68.1%) | 24 (72.7%) |  |
| Education | 16.6 (2.01) | 16.2 (2.16) | 15.3 (2.24) | <0.001*†‡ | 17.0 (1.94) | 16.1 (2.17) | 15.3 (1.86) | <0.001*† |
| PACC 5 | 0.35 (0.93) | 0.08 (0.99) | -0.46 (0.93) | <0.001*†‡ | -0.19 (1.00) | -0.41 (0.96) | -0.67 (0.87) | 0.003*† |
| WMH | 1.94 (2.24) | 1.89 (2.32) | 1.84 (2.02) | 0.946 | 1.69 (2.01) | 2.12 (2.74) | 2.04 (3.39) | 0.278 |
| Amyloid SUVR | 1.08 (0.15) | 1.09 (0.19) | 1.06 (0.13) | 0.425 | 1.07 (0.16) | 1.06 (0.15) | 1.02 (0.11) | 0.487 |
| Tau SUVR | 1.08 (0.19) | 1.07 (0.17) | 1.05 (0.17) | 0.422 | 1.06 (0.13) | 1.02 (0.09) | 1.06 (0.08) | 0.010* |
| GM volume (meta-ROI) | 12646 (1184) | 12453 (1270) | 12016 (1267) | 0.002†‡ | 14249 (1487) | 13930 (1200) | 14312 (1158) | 0.143 |

Note 1: Values are shown as mean (standard deviation) for continuous variables and number (percentage) for categorical variables. P-values for continuous variables were obtained from one-way ANOVA within each sex across ADI categories; p-values for categorical variables were obtained from Fisher’s exact test. For variables with significant overall ANOVA p-values, post-hoc pairwise comparisons with correction for multiple testing were performed (see footnotes).

Note 2: For variables with significant overall p-values, Tukey-adjusted post-hoc pairwise comparisons are indicated by symbols: * Moderate vs Low; † High vs Low; ‡ High vs Moderate (p<0.05).

Note 3: For these comparisons we used the full cohort for the variables that are present in both imaging and full cohort. For the variables only present in imaging cohort only subjects from that cohort were included.

### **Supplementary Table 3.** Demographic data and AD related outcomes stratified by APOE ε4 and ADI level.

|  | **APOE ε4 Carriers** | | | | **APOE ε4 non-Carriers** | | | |
| --- | --- | --- | --- | --- | --- | --- | --- | --- |
|  | **Low** | **Moderate** | **High** | **p** | **Low** | **Moderate** | **High** | **p** |
|  | ***N=295*** | ***N=234*** | ***N=57*** |  | ***N=629*** | ***N=527*** | ***N=138*** |  |
| Age | 67.9 (5.30) | 67.6 (5.08) | 67.5 (5.00) | 0.735 | 68.5 (5.17) | 68.4 (5.15) | 69.1 (4.90) | 0.428 |
| Race: |  |  |  | <0.001 |  |  |  | <0.001 |
| White | 235 (79.7%) | 163 (69.7%) | 19 (33.3%) |  | 502 (79.8%) | 360 (68.3%) | 57 (41.3%) |  |
| African American | 23 (7.80%) | 45 (19.2%) | 32 (56.1%) |  | 27 (4.29%) | 86 (16.3%) | 63 (45.7%) |  |
| Asian | 8 (2.71%) | 1 (0.43%) | 0 (0.00%) |  | 29 (4.61%) | 9 (1.71%) | 1 (0.72%) |  |
| Hispanic | 11 (3.73%) | 9 (3.85%) | 2 (3.51%) |  | 33 (5.25%) | 41 (7.78%) | 10 (7.25%) |  |
| Other or multiple races | 18 (6.10%) | 16 (6.84%) | 4 (7.02%) |  | 38 (6.04%) | 31 (5.88%) | 7 (5.07%) |  |
| Sex: |  |  |  | 0.001 |  |  |  | <0.001 |
| Male | 114 (38.6%) | 68 (29.1%) | 9 (15.8%) |  | 243 (38.6%) | 145 (27.5%) | 24 (17.4%) |  |
| Female | 181 (61.4%) | 166 (70.9%) | 48 (84.2%) |  | 386 (61.4%) | 382 (72.5%) | 114 (82.6%) |  |
| Education | 16.8 (2.07) | 16.2 (2.20) | 15.6 (2.10) | <0.001*† | 16.8 (1.95) | 16.1 (2.15) | 15.1 (2.20) | <0.001*†‡ |
| PACC 5 | 0.04 (1.08) | -0.13 (1.03) | -0.46 (0.93) | 0.003† | 0.19 (0.94) | -0.02 (1.00) | -0.52 (0.92) | <0.001*†‡ |
| WMH | 1.50 (1.79) | 2.06 (2.35) | 2.11 (2.53) | 0.094 | 1.96 (2.26) | 1.93 (2.52) | 1.79 (2.30) | 0.881 |
| Amyloid SUVR | 1.14 (0.19) | 1.19 (0.25) | 1.04 (0.08) | 0.006‡ | 1.05 (0.12) | 1.04 (0.11) | 1.06 (0.14) | 0.369 |
| Tau SUVR | 1.09 (0.18) | 1.09 (0.24) | 1.06 (0.10) | 0.819 | 1.07 (0.15) | 1.04 (0.09) | 1.04 (0.17) | 0.056 |
| GM volume (meta-ROI) | 13436 (1560) | 12983 (1286) | 11932 (1051) | <0.001*†‡ | 13385 (1556) | 12931 (1485) | 12713 (1671) | <0.001*† |

Note: Values are shown as mean (standard deviation) for continuous variables and number (percentage) for categorical variables. P-values for continuous variables were obtained from one-way ANOVA within each sex across ADI categories; p-values for categorical variables were obtained from Fisher’s exact test. For variables with significant overall ANOVA p-values, post-hoc pairwise comparisons with correction for multiple testing were performed (see footnotes).

Note 2: For variables with significant overall p-values, Tukey-adjusted post-hoc pairwise comparisons are indicated by symbols: * Moderate vs Low; † High vs Low; ‡ High vs Moderate (p<0.05).

Note 3: For these comparisons we used the full cohort for the variables that are present in both imaging and full cohort. For the variables only present in imaging cohort only subjects from that cohort were included.

### **Supplementary table 4.** Statistics for interaction model. Outcome: Cognition. ADI continuous.

| **term** | **Sum of Squares** | **Degrees of Freedom** | **F value** | **P-value** |
| --- | --- | --- | --- | --- |
| Intercept | 151.361 | 1 | 196.902 | 0.000 |
| Age | 176.907 | 1 | 230.134 | 0.000 |
| Area Deprivation Index. Nation | 4.547 | 1 | 5.915 | 0.015 |
| Race | 42.171 | 4 | 13.715 | 0.000 |
| Sex | 35.757 | 1 | 46.516 | 0.000 |
| APOE Status | 4.832 | 1 | 6.286 | 0.012 |
| Area Deprivation Index. Nation*Race | 7.073 | 4 | 2.300 | 0.057 |
| Area Deprivation Index. Nation*Sex | 0.113 | 1 | 0.147 | 0.701 |
| Area Deprivation Index. Nation*APOE Status | 0.567 | 1 | 0.737 | 0.391 |
| Residuals | 1,433.647 | 1,865 |  |  |

Abbreviations: APOE = Apolipoprotein E

Note: Sample size N = 1880.

### **Supplementary Table 5.** Statistics for models assessing relationship between ADI continuous and cognition stratified by race and ethnicity.

| **African American** | **Estimate** | **Std. Error** | **t Value** | **P-value** | | **CI Low** | | **CI High** | |
| --- | --- | --- | --- | --- | --- | --- | --- | --- | --- |
| Intercept | 3.572 | 0.783 | 4.561 | 0.000 | | 2.030 | | 5.113 | |
| Area Deprivation Index. Nation | -0.001 | 0.002 | -0.318 | 0.750 | | -0.005 | | 0.004 | |
| Sex: Female | 0.319 | 0.157 | 2.030 | 0.043 | | 0.010 | | 0.629 | |
| Age | -0.067 | 0.011 | -5.855 | 0.000 | | -0.089 | | -0.044 | |
| APOE Status: Non-Carrier | 0.181 | 0.115 | 1.567 | 0.118 | | -0.046 | | 0.408 | |
| **Asian** | **Estimate** | **Std. Error** | **t Value** | **P-value** | | **CI Low** | | **CI High** | |
| Intercept | 2.864 | 1.970 | 1.454 | 0.153 | | -1.109 | | 6.838 | |
| Area Deprivation Index. Nation | -0.005 | 0.009 | -0.542 | 0.590 | | -0.024 | | 0.014 | |
| Sex: Female | 0.315 | 0.304 | 1.038 | 0.305 | | -0.297 | | 0.928 | |
| Age | -0.048 | 0.028 | -1.692 | 0.098 | | -0.104 | | 0.009 | |
| APOE Status: Non-Carrier | 0.154 | 0.365 | 0.422 | 0.675 | | -0.583 | | 0.891 | |
| **Hispanic** | **Estimate** | **Std. Error** | **t Value** | **P-value** | | **CI Low** | | **CI High** | |
| Intercept | 3.436 | 1.090 | 3.151 | 0.002 | | 1.273 | | 5.598 | |
| Area Deprivation Index. Nation | -0.013 | 0.004 | -3.649 | 0.000 | | -0.020 | | -0.006 | |
| Sex: Female | 0.599 | 0.178 | 3.362 | 0.001 | | 0.246 | | 0.953 | |
| Age | -0.050 | 0.016 | -3.120 | 0.002 | | -0.083 | | -0.018 | |
| APOE Status: Non-Carrier | -0.240 | 0.199 | -1.209 | 0.229 | | -0.635 | | 0.154 | |
| **Other** | **Estimate** | **Std. Error** | **t Value** | **P-value** | | **CI Low** | | **CI High** | |
| Intercept | 4.152 | 1.120 | 3.709 | 0.000 | | 1.934 | | 6.371 | |
| Area Deprivation Index. Nation | -0.009 | 0.004 | -2.343 | 0.021 | | -0.017 | | -0.001 | |
| Sex: Female | 0.598 | 0.185 | 3.239 | 0.002 | | 0.232 | | 0.964 | |
| Age | -0.065 | 0.016 | -4.052 | 0.000 | | -0.097 | | -0.033 | |
| APOE Status: Non-Carrier | 0.099 | 0.180 | 0.549 | 0.584 | | -0.258 | | 0.456 | |
| **White** | **Estimate** | **Std. Error** | **t Value** | | **P-value** | | **CI Low** | | **CI High** |
| Intercept | 3.965 | 0.320 | 12.390 | | 0.000 | | 3.338 | | 4.593 |
| Area Deprivation Index. Nation | -0.007 | 0.001 | -6.181 | | 0.000 | | -0.010 | | -0.005 |
| Sex: Female | 0.539 | 0.050 | 10.764 | | 0.000 | | 0.441 | | 0.637 |
| Age | -0.059 | 0.005 | -12.825 | | 0.000 | | -0.068 | | -0.050 |
| APOE Status: Non-Carrier | 0.167 | 0.052 | 3.231 | | 0.001 | | 0.065 | | 0.268 |

### **Supplementary Table 6.** Statistics for interaction model. Outcome: Cognition. ADI categorical

| **term** | **Sum of Squares** | **Degrees of Freedom** | **F value** | **P-value** |
| --- | --- | --- | --- | --- |
| Intercept | 151.424 | 1 | 196.017 | 0.000 |
| Age | 176.384 | 1 | 228.328 | 0.000 |
| Area Deprivation Index | 2.725 | 2 | 1.764 | 0.172 |
| Race | 58.592 | 4 | 18.962 | 0.000 |
| Sex | 61.593 | 1 | 79.731 | 0.000 |
| APOE Status | 4.623 | 1 | 5.985 | 0.015 |
| Area Deprivation Index*Race | 12.526 | 8 | 2.027 | 0.040 |
| Area Deprivation Index*Sex | 1.352 | 2 | 0.875 | 0.417 |
| Area Deprivation Index*APOE Status | 0.753 | 2 | 0.487 | 0.614 |
| Residuals | 1,435.312 | 1,858 |  |  |

Abbreviations: APOE = Apolipoprotein E

Note: Sample size N = 1880.

### **Supplementary Table 7.** Statistics for models assessing relationship between race and ethnicity and cognition stratified by ADI level.

| **ADI: Low** | **Estimate** | **Std. Error** | **t Value** | | **P-value** | | **CI Low** | | **CI High** |
| --- | --- | --- | --- | --- | --- | --- | --- | --- | --- |
| Intercept | 4.004 | 0.382 | 10.469 | | 0.000 | | 3.254 | | 4.755 |
| Sex: Female | 0.531 | 0.059 | 9.006 | | 0.000 | | 0.416 | | 0.647 |
| Age | -0.061 | 0.006 | -11.136 | | 0.000 | | -0.072 | | -0.051 |
| APOE Status: Non-Carrier | 0.153 | 0.062 | 2.486 | | 0.013 | | 0.032 | | 0.274 |
| Race: African American | -1.048 | 0.128 | -8.208 | | 0.000 | | -1.298 | | -0.797 |
| Race: Asian | -0.409 | 0.147 | -2.789 | | 0.005 | | -0.697 | | -0.121 |
| Race: Hispanic | -0.357 | 0.135 | -2.634 | | 0.009 | | -0.622 | | -0.091 |
| Race: Other or multiple races | -0.167 | 0.121 | -1.382 | | 0.167 | | -0.403 | | 0.070 |
| **ADI: Moderate** | **Estimate** | **Std. Error** | **t Value** | **P-value** | | **CI Low** | | **CI High** | |
| Intercept | 3.698 | 0.447 | 8.273 | 0.000 | | 2.820 | | 4.575 | |
| Sex: Female | 0.532 | 0.074 | 7.223 | 0.000 | | 0.387 | | 0.677 | |
| Age | -0.059 | 0.006 | -9.230 | 0.000 | | -0.072 | | -0.047 | |
| APOE Status: Non-Carrier | 0.155 | 0.071 | 2.173 | 0.030 | | 0.015 | | 0.295 | |
| Race: African American | -0.735 | 0.089 | -8.244 | 0.000 | | -0.910 | | -0.560 | |
| Race: Asian | -0.426 | 0.288 | -1.479 | 0.140 | | -0.991 | | 0.139 | |
| Race: Hispanic | -0.515 | 0.134 | -3.838 | 0.000 | | -0.779 | | -0.252 | |
| Race: Other or multiple races | -0.498 | 0.137 | -3.628 | 0.000 | | -0.767 | | -0.229 | |
| **ADI: High** | **Estimate** | **Std. Error** | **t Value** | **P-value** | | **CI Low** | | **CI High** | |
| Intercept | 3.276 | 0.857 | 3.823 | 0.000 | | 1.586 | | 4.967 | |
| Sex: Female | 0.300 | 0.165 | 1.816 | 0.071 | | -0.026 | | 0.626 | |
| Age | -0.054 | 0.012 | -4.414 | 0.000 | | -0.079 | | -0.030 | |
| APOE Status: Non-Carrier | -0.000 | 0.135 | -0.001 | 0.999 | | -0.266 | | 0.266 | |
| Race: African American | -0.523 | 0.133 | -3.921 | 0.000 | | -0.786 | | -0.260 | |
| Race: Asian | 0.252 | 0.850 | 0.296 | 0.767 | | -1.425 | | 1.928 | |
| Race: Hispanic | -0.687 | 0.262 | -2.623 | 0.009 | | -1.203 | | -0.170 | |
| Race: Other or multiple races | 0.144 | 0.273 | 0.526 | 0.600 | | -0.395 | | 0.682 | |

Abbreviation: APOE = Apolipoprotein E; CI = Confidence Interval; Std. = Standard;

Note: Sample size for ADI Low = 924; ADI moderate = 761; ADI High = 195.

### **Supplementary table 8.** Statistics for interaction model. Outcome: Cognition. ADI continuous. Education as covariate.

| **term** | **Sum of Squares** | **Degrees of Freedom** | **F value** | **P-value** |
| --- | --- | --- | --- | --- |
| Intercept | 37.839 | 1 | 52.542 | 0.000 |
| Age | 189.911 | 1 | 263.702 | 0.000 |
| Education | 91.249 | 1 | 126.705 | 0.000 |
| Area Deprivation Index. Nation | 1.295 | 1 | 1.799 | 0.180 |
| Race | 43.016 | 4 | 14.932 | 0.000 |
| Sex | 42.776 | 1 | 59.398 | 0.000 |
| APOE Status | 3.633 | 1 | 5.044 | 0.025 |
| Area Deprivation Index. Nation*Race | 5.316 | 4 | 1.845 | 0.118 |
| Area Deprivation Index. Nation*Sex | 0.600 | 1 | 0.834 | 0.361 |
| Area Deprivation Index. Nation*APOE Status | 0.143 | 1 | 0.199 | 0.656 |
| Residuals | 1,342.398 | 1,864 |  |  |

Abbreviations: APOE = Apolipoprotein E

Note: Sample size N = 1880.

### **Supplementary table 9.** Statistics for interaction model. Outcome: Cognition. ADI categorical. Education as covariate

| **term** | **Sum of Squares** | **Degrees of Freedom** | **F value** | **P-value** |
| --- | --- | --- | --- | --- |
| Intercept | 36.576 | 1 | 50.699 | 0.000 |
| Age | 189.834 | 1 | 263.131 | 0.000 |
| Education | 95.595 | 1 | 132.506 | 0.000 |
| Area Deprivation Index | 0.436 | 2 | 0.302 | 0.739 |
| Race | 54.466 | 4 | 18.874 | 0.000 |
| Sex | 71.105 | 1 | 98.560 | 0.000 |
| APOE Status | 4.435 | 1 | 6.148 | 0.013 |
| Area Deprivation Index*Race | 10.477 | 8 | 1.815 | 0.070 |
| Area Deprivation Index*Sex | 1.827 | 2 | 1.266 | 0.282 |
| Area Deprivation Index*APOE Status | 0.372 | 2 | 0.258 | 0.773 |
| Residuals | 1,339.716 | 1,857 |  |  |

Abbreviations: APOE = Apolipoprotein E

Note: Sample size N = 1880.

### **Supplementary Table 10.** Statistics for models assessing relationship between ADI continuous and cognition stratified by race and ethnicity. Education as covariate

| **African American** | **Estimate** | **Std. Error** | **t Value** | **P-value** | **CI Low** | **CI High** |
| --- | --- | --- | --- | --- | --- | --- |
| Intercept | 1.602 | 0.855 | 1.873 | 0.062 | -0.082 | 3.286 |
| Area Deprivation Index. | 0.002 | 0.002 | 0.749 | 0.455 | -0.003 | 0.006 |
| Sex: Female | 0.266 | 0.152 | 1.756 | 0.080 | -0.032 | 0.565 |
| Age | -0.069 | 0.011 | -6.279 | 0.000 | -0.090 | -0.047 |
| APOE Status: Non-Carrier | 0.167 | 0.111 | 1.502 | 0.134 | -0.052 | 0.385 |
| Education | 0.126 | 0.026 | 4.846 | 0.000 | 0.075 | 0.177 |
| **Asian** | **Estimate** | **Std. Error** | **t Value** | **P-value** | **CI Low** | **CI High** |
| Intercept | 2.726 | 2.410 | 1.131 | 0.264 | -2.137 | 7.590 |
| Area Deprivation Index. | -0.005 | 0.010 | -0.505 | 0.616 | -0.025 | 0.015 |
| Sex: Female | 0.320 | 0.310 | 1.030 | 0.309 | -0.306 | 0.946 |
| Age | -0.047 | 0.028 | -1.669 | 0.103 | -0.105 | 0.010 |
| APOE Status: Non-Carrier | 0.155 | 0.370 | 0.420 | 0.676 | -0.591 | 0.902 |
| Education | 0.007 | 0.073 | 0.102 | 0.919 | -0.140 | 0.155 |
| **Hispanic** | **Estimate** | **Std. Error** | **t Value** | **P-value** | **CI Low** | **CI High** |
| Intercept | 1.544 | 1.182 | 1.306 | 0.194 | -0.801 | 3.889 |
| Area Deprivation Index. | -0.006 | 0.004 | -1.610 | 0.111 | -0.014 | 0.001 |
| Sex: Female | 0.610 | 0.170 | 3.591 | 0.001 | 0.273 | 0.947 |
| Age | -0.057 | 0.016 | -3.668 | 0.000 | -0.088 | -0.026 |
| APOE Status: Non-Carrier | -0.200 | 0.190 | -1.051 | 0.296 | -0.576 | 0.177 |
| Education | 0.130 | 0.039 | 3.354 | 0.001 | 0.053 | 0.208 |
| **Other** | **Estimate** | **Std. Error** | **t Value** | **P-value** | **CI Low** | **CI High** |
| Intercept | 2.940 | 1.210 | 2.429 | 0.017 | 0.541 | 5.339 |
| Area Deprivation Index. | -0.007 | 0.004 | -1.920 | 0.057 | -0.015 | 0.000 |
| Sex: Female | 0.601 | 0.181 | 3.323 | 0.001 | 0.243 | 0.960 |
| Age | -0.071 | 0.016 | -4.454 | 0.000 | -0.102 | -0.039 |
| APOE Status: Non-Carrier | 0.064 | 0.177 | 0.361 | 0.719 | -0.287 | 0.415 |
| Education | 0.095 | 0.040 | 2.368 | 0.020 | 0.016 | 0.175 |
| **White** | **Estimate** | **Std. Error** | **t Value** | **P-value** | **CI Low** | **CI High** |
| Intercept | 2.227 | 0.359 | 6.205 | 0.000 | 1.523 | 2.931 |
| Area Deprivation Index. | -0.005 | 0.001 | -4.176 | 0.000 | -0.007 | -0.003 |
| Sex: Female | 0.565 | 0.048 | 11.645 | 0.000 | 0.470 | 0.660 |
| Age | -0.061 | 0.004 | -13.647 | 0.000 | -0.070 | -0.052 |
| APOE Status: Non-Carrier | 0.174 | 0.050 | 3.477 | 0.001 | 0.076 | 0.272 |
| Education | 0.107 | 0.011 | 9.574 | 0.000 | 0.085 | 0.129 |

### **Supplementary Table 11.** Statistics for models assessing relationship between race and ethnicity and cognition stratified by ADI level. Education included

| **ADI: Low** | **Estimate** | **Std. Error** | **t Value** | | **P-value** | | **CI Low** | | **CI High** |
| --- | --- | --- | --- | --- | --- | --- | --- | --- | --- |
| Intercept | 2.455 | 0.436 | 5.627 | | 0.000 | | 1.598 | | 3.311 |
| Sex: Female | 0.568 | 0.058 | 9.818 | | 0.000 | | 0.454 | | 0.681 |
| Age | -0.063 | 0.005 | -11.678 | | 0.000 | | -0.073 | | -0.052 |
| APOE Status: Non-Carrier | 0.150 | 0.060 | 2.496 | | 0.013 | | 0.032 | | 0.268 |
| Race: African American | -1.012 | 0.125 | -8.116 | | 0.000 | | -1.257 | | -0.767 |
| Race: Asian | -0.406 | 0.143 | -2.841 | | 0.005 | | -0.687 | | -0.126 |
| Race: Hispanic | -0.346 | 0.132 | -2.622 | | 0.009 | | -0.606 | | -0.087 |
| Race: Other or multiple races | -0.161 | 0.118 | -1.373 | | 0.170 | | -0.392 | | 0.069 |
| Education | 0.097 | 0.014 | 6.861 | | 0.000 | | 0.069 | | 0.125 |
| **ADI: Moderate** | **Estimate** | **Std. Error** | **t Value** | | **P-value** | | **CI Low** | | **CI High** |
| Intercept | 1.960 | 0.476 | 4.120 | | 0.000 | | 1.026 | | 2.894 |
| Sex: Female | 0.525 | 0.071 | 7.451 | | 0.000 | | 0.387 | | 0.664 |
| Age | -0.063 | 0.006 | -10.186 | | 0.000 | | -0.075 | | -0.051 |
| APOE Status: Non-Carrier | 0.159 | 0.068 | 2.334 | | 0.020 | | 0.025 | | 0.293 |
| Race: African American | -0.774 | 0.085 | -9.052 | | 0.000 | | -0.942 | | -0.606 |
| Race: Asian | -0.509 | 0.276 | -1.844 | | 0.066 | | -1.050 | | 0.033 |
| Race: Hispanic | -0.379 | 0.130 | -2.926 | | 0.004 | | -0.634 | | -0.125 |
| Race: Other or multiple races | -0.523 | 0.131 | -3.976 | | 0.000 | | -0.780 | | -0.265 |
| Education | 0.122 | 0.015 | 8.355 | | 0.000 | | 0.094 | | 0.151 |
| **ADI: High** | **Estimate** | **Std. Error** | **t Value** | **P-value** | | **CI Low** | | **CI High** | |
| Intercept | 1.679 | 0.910 | 1.845 | 0.067 | | -0.117 | | 3.475 | |
| Sex: Female | 0.298 | 0.159 | 1.881 | 0.062 | | -0.015 | | 0.611 | |
| Age | -0.057 | 0.012 | -4.803 | 0.000 | | -0.080 | | -0.034 | |
| APOE Status: Non-Carrier | 0.045 | 0.130 | 0.347 | 0.729 | | -0.211 | | 0.301 | |
| Race: African American | -0.515 | 0.128 | -4.021 | 0.000 | | -0.767 | | -0.262 | |
| Race: Asian | 0.384 | 0.817 | 0.471 | 0.638 | | -1.227 | | 1.996 | |
| Race: Hispanic | -0.456 | 0.258 | -1.771 | 0.078 | | -0.964 | | 0.052 | |
| Race: Other or multiple races | 0.122 | 0.262 | 0.464 | 0.643 | | -0.396 | | 0.639 | |
| Education | 0.113 | 0.027 | 4.103 | 0.000 | | 0.059 | | 0.167 | |

Abbreviation: APOE = Apolipoprotein E; CI = Confidence Interval; Std. = Standard;

Note: Sample size for ADI Low = 924; ADI moderate = 761; ADI High = 195.

### **Supplementary Table 12.** Statistics for interaction model. Outcome: Amyloid. ADI continuous

| **term** | **Sum of Squares** | **Degrees of Freedom** | **F value** | **P-value** |
| --- | --- | --- | --- | --- |
| Intercept | 2.826 | 1 | 127.973 | 0.000 |
| Age | 0.645 | 1 | 29.202 | 0.000 |
| Area Deprivation Index. Nation | 0.044 | 1 | 1.979 | 0.160 |
| Race | 0.153 | 4 | 1.733 | 0.141 |
| Sex | 0.006 | 1 | 0.272 | 0.602 |
| APOE Status | 0.865 | 1 | 39.179 | 0.000 |
| Area Deprivation Index. Nation*Race | 0.040 | 4 | 0.454 | 0.770 |
| Area Deprivation Index. Nation*Sex | 0.022 | 1 | 0.989 | 0.320 |
| Area Deprivation Index. Nation*APOE Status | 0.027 | 1 | 1.245 | 0.265 |
| Residuals | 18.838 | 853 |  |  |

Abbreviations: APOE = Apolipoprotein E.

Note: Sample size N = 868.

### **Supplementary Table 13.** Statistics for interaction model. Outcome: Amyloid. ADI categorical.

| **term** | **Sum of Squares** | **Degrees of Freedom** | **F value** | **P-value** |
| --- | --- | --- | --- | --- |
| Age | 0.629 | 1 | 29.021 | 0.000 |
| Area Deprivation Index | 0.141 | 2 | 3.242 | 0.040 |
| Race | 0.125 | 4 | 1.440 | 0.219 |
| Sex | 0.028 | 1 | 1.285 | 0.257 |
| APOE Status | 0.856 | 1 | 39.497 | 0.000 |
| Area Deprivation Index*Race | 0.151 | 7 | 0.995 | 0.433 |
| Area Deprivation Index*Sex | 0.034 | 2 | 0.778 | 0.460 |
| Area Deprivation Index*APOE Status | 0.331 | 2 | 7.628 | 0.001 |
| Residuals | 18.364 | 847 |  |  |

Abbreviations: APOE = Apolipoprotein E;

Note: Sample size N = 868.

### **Supplementary Table 14.** Statistics for stratified models by Area Deprivation Index level on Amyloid

| **ADI: Low** | **Estimate** | **Std. Error** | **t Value** | **P-value** | **CI Low** | **CI High** |
| --- | --- | --- | --- | --- | --- | --- |
| Intercept | 0.826 | 0.088 | 9.398 | 0.000 | 0.653 | 0.998 |
| Sex: Female | 0.015 | 0.013 | 1.142 | 0.254 | -0.011 | 0.042 |
| Age | 0.004 | 0.001 | 3.534 | 0.000 | 0.002 | 0.007 |
| APOE Status: Non-Carrier | -0.093 | 0.015 | -6.434 | 0.000 | -0.122 | -0.065 |
| Race: African American | -0.039 | 0.029 | -1.366 | 0.173 | -0.096 | 0.017 |
| Race: Asian | -0.021 | 0.032 | -0.661 | 0.509 | -0.083 | 0.041 |
| Race: Hispanic | -0.026 | 0.029 | -0.871 | 0.384 | -0.083 | 0.032 |
| Race: Other or multiple races | 0.046 | 0.027 | 1.701 | 0.090 | -0.007 | 0.098 |
| **ADI: Moderate** | **Estimate** | **Std. Error** | **t Value** | **P-value** | **CI Low** | **CI High** |
| Intercept | 0.710 | 0.122 | 5.834 | 0.000 | 0.471 | 0.949 |
| Sex: Female | 0.036 | 0.019 | 1.962 | 0.051 | -0.000 | 0.073 |
| Age | 0.007 | 0.002 | 3.857 | 0.000 | 0.003 | 0.010 |
| APOE Status: Non-Carrier | -0.156 | 0.019 | -8.085 | 0.000 | -0.194 | -0.118 |
| Race: African American | -0.035 | 0.024 | -1.478 | 0.140 | -0.082 | 0.012 |
| Race: Asian | 0.074 | 0.065 | 1.137 | 0.256 | -0.054 | 0.203 |
| Race: Hispanic | -0.014 | 0.031 | -0.446 | 0.656 | -0.074 | 0.047 |
| Race: Other or multiple races | 0.093 | 0.035 | 2.674 | 0.008 | 0.025 | 0.161 |
| **ADI: High** | **Estimate** | **Std. Error** | **t Value** | **P-value** | **CI Low** | **CI High** |
| Intercept | 0.748 | 0.196 | 3.813 | 0.000 | 0.357 | 1.139 |
| Sex: Female | 0.063 | 0.037 | 1.692 | 0.095 | -0.011 | 0.137 |
| Age | 0.004 | 0.003 | 1.433 | 0.156 | -0.002 | 0.010 |
| APOE Status: Non-Carrier | 0.002 | 0.033 | 0.053 | 0.958 | -0.065 | 0.068 |
| Race: African American | -0.049 | 0.033 | -1.502 | 0.138 | -0.115 | 0.016 |
| Race: Hispanic | -0.001 | 0.052 | -0.022 | 0.982 | -0.105 | 0.103 |
| Race: Other or multiple races | -0.075 | 0.061 | -1.231 | 0.223 | -0.197 | 0.047 |

Abbreviations: APOE = Apolipoprotein E; Std. = Standard; CI = Confidence Interval

Note: Sample size for ADI low = 463; ADI moderate = 329; ADI High = 76.

### **Supplementary Table 15.** Statistics for interaction model. Outcome: Amyloid. ADI categorical. Education included in model

| **term** | **Sum of Squares** | **Degrees of Freedom** | **F value** | **P-value** |
| --- | --- | --- | --- | --- |
| Age | 0.639 | 1 | 29.447 | 0.000 |
| Education | 0.012 | 1 | 0.554 | 0.457 |
| Area Deprivation Index | 0.137 | 2 | 3.150 | 0.043 |
| Race | 0.127 | 4 | 1.466 | 0.211 |
| Sex | 0.026 | 1 | 1.204 | 0.273 |
| APOE Status | 0.856 | 1 | 39.449 | 0.000 |
| Area Deprivation Index*Race | 0.152 | 7 | 1.002 | 0.428 |
| Area Deprivation Index*Sex | 0.035 | 2 | 0.806 | 0.447 |
| Area Deprivation Index*APOE Status | 0.327 | 2 | 7.546 | 0.001 |
| Residuals | 18.352 | 846 |  |  |

Abbreviations: APOE = Apolipoprotein E;

Note: Sample size N = 868.

### **Supplementary Table 16.** Statistics for stratified models by Area Deprivation Index on Amyloid. Education included in model

| **ADI: Low** | **Estimate** | **Std. Error** | **t Value** | **P-value** | **CI Low** | **CI High** |
| --- | --- | --- | --- | --- | --- | --- |
| Intercept | 0.827 | 0.102 | 8.085 | 0.000 | 0.626 | 1.028 |
| Sex: Female | 0.015 | 0.013 | 1.137 | 0.256 | -0.011 | 0.042 |
| Age | 0.004 | 0.001 | 3.525 | 0.000 | 0.002 | 0.007 |
| APOE Status: Non-Carrier | -0.093 | 0.015 | -6.426 | 0.000 | -0.122 | -0.065 |
| Race: African American | -0.039 | 0.029 | -1.363 | 0.173 | -0.096 | 0.017 |
| Race: Asian | -0.021 | 0.032 | -0.660 | 0.510 | -0.083 | 0.041 |
| Race: Hispanic | -0.025 | 0.029 | -0.869 | 0.386 | -0.083 | 0.032 |
| Race: Other or multiple races | 0.046 | 0.027 | 1.699 | 0.090 | -0.007 | 0.098 |
| Education | -0.000 | 0.003 | -0.024 | 0.981 | -0.007 | 0.007 |
| **ADI: Moderate** | **Estimate** | **Std. Error** | **t Value** | **P-value** | **CI Low** | **CI High** |
| Intercept | 0.764 | 0.133 | 5.754 | 0.000 | 0.503 | 1.026 |
| Sex: Female | 0.037 | 0.019 | 1.996 | 0.047 | 0.001 | 0.074 |
| Age | 0.007 | 0.002 | 3.936 | 0.000 | 0.003 | 0.010 |
| APOE Status: Non-Carrier | -0.156 | 0.019 | -8.094 | 0.000 | -0.194 | -0.118 |
| Race: African American | -0.034 | 0.024 | -1.438 | 0.152 | -0.081 | 0.013 |
| Race: Asian | 0.077 | 0.065 | 1.175 | 0.241 | -0.052 | 0.205 |
| Race: Hispanic | -0.018 | 0.031 | -0.596 | 0.552 | -0.079 | 0.043 |
| Race: Other or multiple races | 0.095 | 0.035 | 2.738 | 0.007 | 0.027 | 0.164 |
| Education | -0.004 | 0.004 | -1.022 | 0.308 | -0.012 | 0.004 |
| **ADI: High** | **Estimate** | **Std. Error** | **t Value** | **P-value** | **CI Low** | **CI High** |
| Intercept | 0.763 | 0.225 | 3.394 | 0.001 | 0.315 | 1.212 |
| Sex: Female | 0.063 | 0.037 | 1.675 | 0.098 | -0.012 | 0.137 |
| Age | 0.004 | 0.003 | 1.428 | 0.158 | -0.002 | 0.010 |
| APOE Status: Non-Carrier | 0.001 | 0.034 | 0.033 | 0.974 | -0.066 | 0.069 |
| Race: African American | -0.050 | 0.034 | -1.493 | 0.140 | -0.117 | 0.017 |
| Race: Hispanic | -0.004 | 0.055 | -0.065 | 0.948 | -0.114 | 0.106 |
| Race: Other or multiple races | -0.076 | 0.062 | -1.226 | 0.224 | -0.199 | 0.047 |
| Education | -0.001 | 0.007 | -0.142 | 0.887 | -0.015 | 0.013 |

Abbreviations: APOE = Apolipoprotein E; Std. = Standard; CI = Confidence Interval.

Note: Sample size for ADI low = 463; ADI moderate = 329; ADI High = 76.

### **Supplementary Table 17.** Statistics for interaction model. Outcome: Amyloid. ADI categorical (centiloids).

| **term** | **Sum of Squares** | **Degrees of Freedom** | **F value** | **P-value** |
| --- | --- | --- | --- | --- |
| Age | 15,672.846 | 1 | 29.213 | 0.000 |
| Education | 235.944 | 1 | 0.440 | 0.507 |
| Area Deprivation Index | 3,309.434 | 2 | 3.084 | 0.046 |
| Race | 3,280.221 | 4 | 1.529 | 0.192 |
| Sex | 690.244 | 1 | 1.287 | 0.257 |
| APOE Status | 21,170.050 | 1 | 39.459 | 0.000 |
| Area Deprivation Index*Race | 4,070.109 | 7 | 1.084 | 0.372 |
| Area Deprivation Index*Sex | 867.795 | 2 | 0.809 | 0.446 |
| Area Deprivation Index*APOE Status | 8,192.279 | 2 | 7.635 | 0.001 |
| Residuals | 453,883.179 | 846 |  |  |

### **Supplementary Table 18.** Statistics for interaction model. Outcome: Amyloid. ADI continuous (centiloids).

| **term** | **Sum of Squares** | **Degrees of Freedom** | **F value** | **P-value** |
| --- | --- | --- | --- | --- |
| Intercept | 2,971.468 | 1 | 5.437 | 0.020 |
| Age | 15,878.341 | 1 | 29.055 | 0.000 |
| Area Deprivation Index. Nation | 1,069.447 | 1 | 1.957 | 0.162 |
| Race | 4,061.695 | 4 | 1.858 | 0.116 |
| Sex | 162.872 | 1 | 0.298 | 0.585 |
| APOE Status | 21,317.706 | 1 | 39.008 | 0.000 |
| Area Deprivation Index. Nation*Race | 1,042.261 | 4 | 0.477 | 0.753 |
| Area Deprivation Index. Nation*Sex | 549.886 | 1 | 1.006 | 0.316 |
| Area Deprivation Index. Nation*APOE Status | 635.661 | 1 | 1.163 | 0.281 |
| Residuals | 466,156.373 | 853 |  |  |

**Supplementary Table 19.** Statistics for stratified models by Area Deprivation Index on Amyloid.

| **ADI: Low** | **Estimate** | **Std. Error** | **t Value** | **P-value** | **CI Low** | **CI High** |
| --- | --- | --- | --- | --- | --- | --- |
| Intercept | -21.657 | 13.821 | -1.567 | 0.118 | -48.818 | 5.504 |
| Sex: Female | 2.478 | 2.109 | 1.175 | 0.241 | -1.667 | 6.623 |
| Age | 0.692 | 0.199 | 3.486 | 0.001 | 0.302 | 1.082 |
| APOE Status: Non-Carrier | -14.677 | 2.282 | -6.431 | 0.000 | -19.161 | -10.192 |
| Race: African American | -6.362 | 4.536 | -1.403 | 0.161 | -15.275 | 2.552 |
| Race: Asian | -3.141 | 4.974 | -0.632 | 0.528 | -12.915 | 6.633 |
| Race: Hispanic | -4.199 | 4.609 | -0.911 | 0.363 | -13.257 | 4.859 |
| Race: Other or multiple races | 7.347 | 4.220 | 1.741 | 0.082 | -0.945 | 15.640 |
| **ADI: Moderate** | **Estimate** | **Std. Error** | **t Value** | **P-value** | **CI Low** | **CI High** |
| Intercept | -41.291 | 19.123 | -2.159 | 0.032 | -78.913 | -3.669 |
| Sex: Female | 5.910 | 2.918 | 2.026 | 0.044 | 0.170 | 11.651 |
| Age | 1.061 | 0.272 | 3.901 | 0.000 | 0.526 | 1.595 |
| APOE Status: Non-Carrier | -24.631 | 3.026 | -8.139 | 0.000 | -30.586 | -18.677 |
| Race: African American | -5.809 | 3.746 | -1.551 | 0.122 | -13.178 | 1.561 |
| Race: Asian | 11.913 | 10.267 | 1.160 | 0.247 | -8.285 | 32.111 |
| Race: Hispanic | -2.291 | 4.814 | -0.476 | 0.634 | -11.762 | 7.180 |
| Race: Other or multiple races | 14.644 | 5.448 | 2.688 | 0.008 | 3.927 | 25.362 |
| **ADI: High** | **Estimate** | **Std. Error** | **t Value** | **P-value** | **CI Low** | **CI High** |
| Intercept | -33.305 | 30.922 | -1.077 | 0.285 | -94.993 | 28.382 |
| Sex: Female | 9.801 | 5.844 | 1.677 | 0.098 | -1.858 | 21.460 |
| Age | 0.631 | 0.451 | 1.399 | 0.166 | -0.269 | 1.530 |
| APOE Status: Non-Carrier | 0.213 | 5.239 | 0.041 | 0.968 | -10.239 | 10.665 |
| Race: African American | -7.685 | 5.168 | -1.487 | 0.142 | -17.995 | 2.626 |
| Race: Hispanic | -0.242 | 8.205 | -0.029 | 0.977 | -16.610 | 16.126 |
| Race: Other or multiple races | -13.248 | 9.651 | -1.373 | 0.174 | -32.501 | 6.006 |

**Supplementary Table 20.** Statistics for stratified models by Area Deprivation Index on Amyloid (centiloids). Education included in model

| **term** | **Sum of Squares** | **Degrees of Freedom** | **F value** | **P-value** |
| --- | --- | --- | --- | --- |
| Age | 15,672.846 | 1 | 29.213 | 0.000 |
| Education | 235.944 | 1 | 0.440 | 0.507 |
| Area Deprivation Index | 3,309.434 | 2 | 3.084 | 0.046 |
| Race | 3,280.221 | 4 | 1.529 | 0.192 |
| Sex | 690.244 | 1 | 1.287 | 0.257 |
| APOE Status | 21,170.050 | 1 | 39.459 | 0.000 |
| Area Deprivation Index*Race | 4,070.109 | 7 | 1.084 | 0.372 |
| Area Deprivation Index*Sex | 867.795 | 2 | 0.809 | 0.446 |
| Area Deprivation Index*APOE Status | 8,192.279 | 2 | 7.635 | 0.001 |
| Residuals | 453,883.179 | 846 |  |  |

### **Supplementary Table 21.** Statistics for interaction model. Outcome: Tau. ADI continuous.

| **term** | **Sum of Squares** | **Degrees of Freedom** | **F value** | **P-value** |
| --- | --- | --- | --- | --- |
| Intercept | 4.044 | 1 | 468.116 | 0.000 |
| Age | 0.020 | 1 | 2.294 | 0.130 |
| Area Deprivation Index. Nation | 0.000 | 1 | 0.004 | 0.948 |
| Race | 0.064 | 4 | 1.846 | 0.118 |
| Sex | 0.002 | 1 | 0.225 | 0.635 |
| APOE Status | 0.002 | 1 | 0.274 | 0.601 |
| Area Deprivation Index. Nation*Race | 0.059 | 4 | 1.702 | 0.147 |
| Area Deprivation Index. Nation*Sex | 0.001 | 1 | 0.118 | 0.732 |
| Area Deprivation Index. Nation*APOE Status | 0.021 | 1 | 2.405 | 0.121 |
| Residuals | 6.919 | 801 |  |  |

Abbreviations: APOE = Apolipoprotein E.

Note: Sample size N = 816.

### **Supplementary Table 22.** Statistics for interaction model. Outcome: Tau. ADI categorical

| **term** | **Sum of Squares** | **Degrees of Freedom** | **F value** | **P-value** |
| --- | --- | --- | --- | --- |
| Age | 0.022 | 1 | 2.533 | 0.112 |
| Area Deprivation Index | 0.047 | 2 | 2.735 | 0.065 |
| Race | 0.048 | 4 | 1.391 | 0.235 |
| Sex | 0.000 | 1 | 0.000 | 0.987 |
| APOE Status | 0.000 | 1 | 0.018 | 0.892 |
| Area Deprivation Index*Race | 0.058 | 7 | 0.966 | 0.455 |
| Area Deprivation Index*Sex | 0.073 | 2 | 4.223 | 0.015 |
| Area Deprivation Index*APOE Status | 0.048 | 2 | 2.809 | 0.061 |
| Residuals | 6.828 | 795 |  |  |

Abbreviations: APOE = Apolipoprotein E;

Note: Sample size N = 816.

### **Supplementary Table 23.** Statistics for stratified models by Area Deprivation Index on Tau

| **ADI: Low** | **Estimate** | **Std. Error** | **t Value** | **P-value** | **CI Low** | **CI High** |
| --- | --- | --- | --- | --- | --- | --- |
| Intercept | 1.014 | 0.058 | 17.585 | 0.000 | 0.901 | 1.127 |
| Sex: Female | -0.000 | 0.009 | -0.011 | 0.991 | -0.017 | 0.017 |
| Age | 0.001 | 0.001 | 0.646 | 0.519 | -0.001 | 0.002 |
| APOE Status: Non-Carrier | -0.001 | 0.010 | -0.110 | 0.913 | -0.020 | 0.018 |
| Race: African American | -0.020 | 0.019 | -1.008 | 0.314 | -0.058 | 0.019 |
| Race: Asian | 0.001 | 0.021 | 0.054 | 0.957 | -0.039 | 0.042 |
| Race: Hispanic | -0.043 | 0.019 | -2.281 | 0.023 | -0.079 | -0.006 |
| Race: Other or multiple races | -0.006 | 0.018 | -0.302 | 0.763 | -0.041 | 0.030 |
| **ADI: Moderate** | **Estimate** | **Std. Error** | **t Value** | **P-value** | **CI Low** | **CI High** |
| Intercept | 0.919 | 0.074 | 12.343 | 0.000 | 0.773 | 1.066 |
| Sex: Female | 0.032 | 0.011 | 2.842 | 0.005 | 0.010 | 0.055 |
| Age | 0.002 | 0.001 | 1.580 | 0.115 | -0.000 | 0.004 |
| APOE Status: Non-Carrier | -0.012 | 0.012 | -1.028 | 0.305 | -0.036 | 0.011 |
| Race: African American | -0.025 | 0.015 | -1.706 | 0.089 | -0.054 | 0.004 |
| Race: Asian | 0.005 | 0.039 | 0.119 | 0.905 | -0.072 | 0.082 |
| Race: Hispanic | -0.007 | 0.019 | -0.396 | 0.693 | -0.044 | 0.029 |
| Race: Other or multiple races | -0.020 | 0.021 | -0.945 | 0.346 | -0.062 | 0.022 |
| **ADI: High** | **Estimate** | **Std. Error** | **t Value** | **P-value** | **CI Low** | **CI High** |
| Intercept | 1.042 | 0.153 | 6.794 | 0.000 | 0.736 | 1.348 |
| Sex: Female | -0.042 | 0.029 | -1.456 | 0.150 | -0.101 | 0.016 |
| Age | 0.001 | 0.002 | 0.502 | 0.618 | -0.003 | 0.006 |
| APOE Status: Non-Carrier | -0.065 | 0.026 | -2.474 | 0.016 | -0.118 | -0.013 |
| Race: African American | -0.021 | 0.026 | -0.829 | 0.410 | -0.073 | 0.030 |
| Race: Hispanic | 0.048 | 0.043 | 1.101 | 0.275 | -0.039 | 0.134 |
| Race: Other or multiple races | -0.065 | 0.048 | -1.356 | 0.180 | -0.161 | 0.031 |

Abbreviations: APOE = Apolipoprotein E; Std. = Standard; CI = Confidence Interval

Note: Sample size ADI Low = 431; ADI Moderate = 312; ADI High = 73.

### **Supplementary Table 24.** Statistics for interaction model. Outcome: Tau. ADI categorical. Education included in model

| **term** | **Sum of Squares** | **Degrees of Freedom** | **F value** | **P-value** |
| --- | --- | --- | --- | --- |
| Age | 0.020 | 1 | 2.322 | 0.128 |
| Education | 0.004 | 1 | 0.415 | 0.520 |
| Area Deprivation Index | 0.046 | 2 | 2.685 | 0.069 |
| Race | 0.048 | 4 | 1.395 | 0.234 |
| Sex | 0.000 | 1 | 0.002 | 0.966 |
| APOE Status | 0.000 | 1 | 0.020 | 0.887 |
| Area Deprivation Index*Race | 0.060 | 7 | 1.003 | 0.427 |
| Area Deprivation Index*Sex | 0.072 | 2 | 4.163 | 0.016 |
| Area Deprivation Index*APOE Status | 0.047 | 2 | 2.728 | 0.066 |
| Residuals | 6.824 | 794 |  |  |

Abbreviations: APOE = Apolipoprotein E;

Note: Sample size N = 816.

### **Supplementary Table 25.** Statistics for stratified models by Area Deprivation Index on Tau. Education included in model

| **ADI: Low** | **Estimate** | **Std. Error** | **t Value** | **P-value** | **CI Low** | **CI High** |
| --- | --- | --- | --- | --- | --- | --- |
| Intercept | 0.985 | 0.066 | 14.881 | 0.000 | 0.855 | 1.116 |
| Sex: Female | 0.000 | 0.009 | 0.040 | 0.968 | -0.017 | 0.018 |
| Age | 0.000 | 0.001 | 0.550 | 0.582 | -0.001 | 0.002 |
| APOE Status: Non-Carrier | -0.001 | 0.010 | -0.123 | 0.902 | -0.020 | 0.018 |
| Race: African American | -0.018 | 0.019 | -0.945 | 0.345 | -0.057 | 0.020 |
| Race: Asian | 0.001 | 0.021 | 0.057 | 0.955 | -0.039 | 0.042 |
| Race: Hispanic | -0.043 | 0.019 | -2.309 | 0.021 | -0.080 | -0.006 |
| Race: Other or multiple races | -0.006 | 0.018 | -0.329 | 0.742 | -0.042 | 0.030 |
| Education | 0.002 | 0.002 | 0.879 | 0.380 | -0.002 | 0.007 |
| **ADI: Moderate** | **Estimate** | **Std. Error** | **t Value** | **P-value** | **CI Low** | **CI High** |
| Intercept | 0.926 | 0.082 | 11.318 | 0.000 | 0.765 | 1.087 |
| Sex: Female | 0.032 | 0.011 | 2.841 | 0.005 | 0.010 | 0.055 |
| Age | 0.002 | 0.001 | 1.589 | 0.113 | -0.000 | 0.004 |
| APOE Status: Non-Carrier | -0.012 | 0.012 | -1.031 | 0.303 | -0.036 | 0.011 |
| Race: African American | -0.025 | 0.015 | -1.695 | 0.091 | -0.054 | 0.004 |
| Race: Asian | 0.005 | 0.039 | 0.126 | 0.900 | -0.072 | 0.082 |
| Race: Hispanic | -0.008 | 0.019 | -0.420 | 0.675 | -0.045 | 0.029 |
| Race: Other or multiple races | -0.020 | 0.021 | -0.929 | 0.354 | -0.062 | 0.022 |
| Education | -0.000 | 0.002 | -0.198 | 0.843 | -0.005 | 0.004 |
| **ADI: High** | **Estimate** | **Std. Error** | **t Value** | **P-value** | **CI Low** | **CI High** |
| Intercept | 0.992 | 0.175 | 5.675 | 0.000 | 0.643 | 1.341 |
| Sex: Female | -0.042 | 0.029 | -1.439 | 0.155 | -0.101 | 0.016 |
| Age | 0.001 | 0.002 | 0.455 | 0.651 | -0.003 | 0.006 |
| APOE Status: Non-Carrier | -0.063 | 0.027 | -2.342 | 0.022 | -0.116 | -0.009 |
| Race: African American | -0.018 | 0.026 | -0.699 | 0.487 | -0.071 | 0.034 |
| Race: Hispanic | 0.057 | 0.046 | 1.231 | 0.223 | -0.035 | 0.148 |
| Race: Other or multiple races | -0.064 | 0.048 | -1.320 | 0.191 | -0.160 | 0.033 |
| Education | 0.003 | 0.006 | 0.605 | 0.547 | -0.008 | 0.015 |

Abbreviations: APOE = Apolipoprotein E; Std. = Standard; CI = Confidence Interval.

Note: Sample size ADI Low = 431; ADI Moderate = 312; ADI High = 73.

**Supplementary Table 26.** Statistics for interaction model. Outcome: WMH. ADI continuous

| **term** | **Sum of Squares** | **Degrees of Freedom** | **F value** | **P-value** |
| --- | --- | --- | --- | --- |
| Intercept | 25.223 | 1 | 75.259 | 0.000 |
| Age | 47.734 | 1 | 142.427 | 0.000 |
| Area Deprivation Index. Nation | 1.003 | 1 | 2.993 | 0.084 |
| Race | 3.145 | 4 | 2.346 | 0.053 |
| Sex | 1.707 | 1 | 5.093 | 0.024 |
| APOE Status | 1.411 | 1 | 4.211 | 0.040 |
| Area Deprivation Index. Nation*Race | 2.752 | 4 | 2.053 | 0.085 |
| Area Deprivation Index. Nation*Sex | 0.079 | 1 | 0.235 | 0.628 |
| Area Deprivation Index. Nation*APOE Status | 2.029 | 1 | 6.055 | 0.014 |
| Residuals | 273.146 | 815 |  |  |

Abbreviations: APOE = Apolipoprotein E;

Note: Sample size N = 830.

### **Supplementary Table 27.** Statistics for interaction model. Outcome: WMH. ADI categorical

| **term** | **Sum of Squares** | **Degrees of Freedom** | **F value** | **P-value** |
| --- | --- | --- | --- | --- |
| Age | 46.938 | 1 | 139.059 | 0.000 |
| Area Deprivation Index | 1.052 | 2 | 1.558 | 0.211 |
| Race | 2.250 | 4 | 1.667 | 0.156 |
| Sex | 3.030 | 1 | 8.977 | 0.003 |
| APOE Status | 0.861 | 1 | 2.550 | 0.111 |
| Area Deprivation Index*Race | 2.251 | 7 | 0.953 | 0.465 |
| Area Deprivation Index*Sex | 0.145 | 2 | 0.214 | 0.807 |
| Area Deprivation Index*APOE Status | 2.498 | 2 | 3.700 | 0.025 |
| Residuals | 273.070 | 809 |  |  |

Abbreviations: APOE = Apolipoprotein E;

Note: Sample size N = 830.

### **Supplementary Table 28.** Statistics for stratified models by APOE status on WMH. ADI continuous.

| **APOE ε4 Carriers** | **Estimate** | **Std. Error** | **t Value** | **P-value** | **CI Low** | **CI High** |
| --- | --- | --- | --- | --- | --- | --- |
| Intercept | -2.769 | 0.468 | -5.919 | 0.000 | -3.690 | -1.847 |
| Area Deprivation Index. Nation | 0.004 | 0.002 | 2.068 | 0.040 | 0.000 | 0.007 |
| Sex: Female | 0.086 | 0.074 | 1.155 | 0.249 | -0.061 | 0.233 |
| Age | 0.052 | 0.007 | 7.805 | 0.000 | 0.039 | 0.065 |
| Race: African American | -0.062 | 0.103 | -0.596 | 0.552 | -0.265 | 0.142 |
| Race: Asian | -0.291 | 0.254 | -1.145 | 0.253 | -0.791 | 0.209 |
| Race: Hispanic | -0.059 | 0.151 | -0.389 | 0.698 | -0.357 | 0.239 |
| Race: Other or multiple races | -0.049 | 0.135 | -0.365 | 0.715 | -0.314 | 0.216 |
| **APOE ε4 non-Carriers** | **Estimate** | **Std. Error** | **t Value** | **P-value** | **CI Low** | **CI High** |
| Intercept | -2.110 | 0.340 | -6.210 | 0.000 | -2.778 | -1.443 |
| Area Deprivation Index. Nation | -0.001 | 0.001 | -0.895 | 0.371 | -0.004 | 0.001 |
| Sex: Female | 0.175 | 0.052 | 3.388 | 0.001 | 0.074 | 0.276 |
| Age | 0.044 | 0.005 | 8.982 | 0.000 | 0.034 | 0.054 |
| Race: African American | -0.122 | 0.084 | -1.461 | 0.145 | -0.286 | 0.042 |
| Race: Asian | -0.162 | 0.131 | -1.235 | 0.217 | -0.419 | 0.096 |
| Race: Hispanic | -0.066 | 0.093 | -0.711 | 0.477 | -0.250 | 0.117 |
| Race: Other or multiple races | -0.119 | 0.102 | -1.168 | 0.243 | -0.319 | 0.081 |

### **Supplementary Table 29.** Statistics for stratified models by Area Deprivation Index on WMH.

| **ADI: Low** | **Estimate** | **Std. Error** | **t Value** | **P-value** | **CI Low** | **CI High** |
| --- | --- | --- | --- | --- | --- | --- |
| Intercept | -2.357 | 0.362 | -6.514 | 0.000 | -3.068 | -1.646 |
| Sex: Female | 0.165 | 0.055 | 3.007 | 0.003 | 0.057 | 0.273 |
| Age | 0.046 | 0.005 | 8.884 | 0.000 | 0.036 | 0.056 |
| APOE Status: Non-Carrier | 0.095 | 0.059 | 1.606 | 0.109 | -0.021 | 0.211 |
| Race: African American | 0.052 | 0.119 | 0.443 | 0.658 | -0.181 | 0.285 |
| Race: Asian | -0.224 | 0.128 | -1.755 | 0.080 | -0.476 | 0.027 |
| Race: Hispanic | -0.190 | 0.123 | -1.540 | 0.124 | -0.431 | 0.052 |
| Race: Other or multiple races | -0.141 | 0.110 | -1.277 | 0.202 | -0.357 | 0.076 |
| **ADI: Moderate** | **Estimate** | **Std. Error** | **t Value** | **P-value** | **CI Low** | **CI High** |
| Intercept | -2.423 | 0.480 | -5.050 | 0.000 | -3.368 | -1.479 |
| Sex: Female | 0.111 | 0.073 | 1.510 | 0.132 | -0.034 | 0.255 |
| Age | 0.049 | 0.007 | 7.190 | 0.000 | 0.036 | 0.063 |
| APOE Status: Non-Carrier | -0.067 | 0.075 | -0.898 | 0.370 | -0.215 | 0.080 |
| Race: African American | -0.104 | 0.094 | -1.103 | 0.271 | -0.290 | 0.082 |
| Race: Asian | -0.008 | 0.271 | -0.028 | 0.978 | -0.541 | 0.526 |
| Race: Hispanic | -0.031 | 0.118 | -0.264 | 0.792 | -0.264 | 0.201 |
| Race: Other or multiple races | -0.052 | 0.135 | -0.385 | 0.700 | -0.317 | 0.213 |
| **ADI: High** | **Estimate** | **Std. Error** | **t Value** | **P-value** | **CI Low** | **CI High** |
| Intercept | -1.414 | 0.871 | -1.624 | 0.109 | -3.152 | 0.324 |
| Sex: Female | 0.108 | 0.160 | 0.674 | 0.503 | -0.212 | 0.427 |
| Age | 0.036 | 0.013 | 2.885 | 0.005 | 0.011 | 0.062 |
| APOE Status: Non-Carrier | -0.293 | 0.144 | -2.033 | 0.046 | -0.581 | -0.005 |
| Race: African American | -0.130 | 0.142 | -0.911 | 0.366 | -0.414 | 0.155 |
| Race: Hispanic | 0.296 | 0.224 | 1.317 | 0.192 | -0.152 | 0.743 |
| Race: Other or multiple races | 0.094 | 0.290 | 0.323 | 0.747 | -0.485 | 0.672 |

Abbreviations: APOE = Apolipoprotein E; Std. = Standard; CI = Confidence Interval

Note: Sample size ADI Low = 425; ADI Moderate = 302; ADI High = 73.

**Supplementary Table 30.** Statistics for interaction model. Outcome: WMH. ADI continuous. Education included in model.

| **term** | **Sum of Squares** | **Degrees of Freedom** | **F value** | **P-value** |
| --- | --- | --- | --- | --- |
| Intercept | 19.742 | 1 | 58.833 | 0.000 |
| Age | 47.466 | 1 | 141.454 | 0.000 |
| Education | 0.000 | 1 | 0.001 | 0.974 |
| Area Deprivation Index. Nation | 0.999 | 1 | 2.976 | 0.085 |
| Race | 3.140 | 4 | 2.340 | 0.054 |
| Sex | 1.704 | 1 | 5.079 | 0.024 |
| APOE Status | 1.408 | 1 | 4.195 | 0.041 |
| Area Deprivation Index. Nation*Race | 2.741 | 4 | 2.042 | 0.087 |
| Area Deprivation Index. Nation*Sex | 0.079 | 1 | 0.235 | 0.628 |
| Area Deprivation Index. Nation*APOE Status | 2.024 | 1 | 6.031 | 0.014 |
| Residuals | 273.146 | 814 |  |  |

### **Supplementary Table 31.** Statistics for stratified models by APOE status on WMH. ADI continuous. Education included in model

| **APOE ε4 Carriers** | **Estimate** | **Std. Error** | **t Value** | **P-value** | **CI Low** | **CI High** |
| --- | --- | --- | --- | --- | --- | --- |
| Intercept | -2.575 | 0.519 | -4.966 | 0.000 | -3.597 | -1.554 |
| Area Deprivation Index. Nation | 0.003 | 0.002 | 1.937 | 0.054 | -0.000 | 0.007 |
| Sex: Female | 0.089 | 0.075 | 1.193 | 0.234 | -0.058 | 0.236 |
| Age | 0.053 | 0.007 | 7.846 | 0.000 | 0.039 | 0.066 |
| Race: African American | -0.071 | 0.104 | -0.679 | 0.498 | -0.275 | 0.134 |
| Race: Asian | -0.297 | 0.254 | -1.167 | 0.244 | -0.798 | 0.204 |
| Race: Hispanic | -0.069 | 0.152 | -0.455 | 0.650 | -0.368 | 0.230 |
| Race: Other or multiple races | -0.052 | 0.135 | -0.383 | 0.702 | -0.317 | 0.214 |
| Education | -0.015 | 0.017 | -0.866 | 0.387 | -0.048 | 0.019 |
| **APOE ε4 non-Carriers** | **Estimate** | **Std. Error** | **t Value** | **P-value** | **CI Low** | **CI High** |
| Intercept | -2.173 | 0.393 | -5.524 | 0.000 | -2.945 | -1.400 |
| Area Deprivation Index. Nation | -0.001 | 0.001 | -0.784 | 0.434 | -0.003 | 0.001 |
| Sex: Female | 0.176 | 0.052 | 3.398 | 0.001 | 0.074 | 0.278 |
| Age | 0.044 | 0.005 | 8.952 | 0.000 | 0.034 | 0.053 |
| Race: African American | -0.123 | 0.084 | -1.472 | 0.142 | -0.288 | 0.041 |
| Race: Asian | -0.163 | 0.131 | -1.241 | 0.215 | -0.420 | 0.095 |
| Race: Hispanic | -0.064 | 0.094 | -0.680 | 0.497 | -0.248 | 0.120 |
| Race: Other or multiple races | -0.121 | 0.102 | -1.183 | 0.237 | -0.322 | 0.080 |
| Education | 0.004 | 0.012 | 0.317 | 0.752 | -0.020 | 0.028 |

### **Supplementary Table 32.** Statistics for interaction model. Outcome: WMH. ADI categorical. Education included in model

| **term** | **Sum of Squares** | **Degrees of Freedom** | **F value** | **P-value** |
| --- | --- | --- | --- | --- |
| Age | 46.714 | 1 | 138.224 | 0.000 |
| Education | 0.000 | 1 | 0.000 | 0.991 |
| Area Deprivation Index | 1.046 | 2 | 1.548 | 0.213 |
| Race | 2.246 | 4 | 1.662 | 0.157 |
| Sex | 3.025 | 1 | 8.952 | 0.003 |
| APOE Status | 0.861 | 1 | 2.547 | 0.111 |
| Area Deprivation Index*Race | 2.233 | 7 | 0.944 | 0.471 |
| Area Deprivation Index*Sex | 0.144 | 2 | 0.214 | 0.808 |
| Area Deprivation Index*APOE Status | 2.496 | 2 | 3.693 | 0.025 |
| Residuals | 273.070 | 808 |  |  |

Abbreviations: APOE = Apolipoprotein E;

Note: Sample size N = 830.

### **Supplementary Table 33.** Statistics for stratified models by Area Deprivation Index on WMH. Education included in model

| **ADI: Low** | **Estimate** | **Std. Error** | **t Value** | **P-value** | **CI Low** | **CI High** |
| --- | --- | --- | --- | --- | --- | --- |
| Intercept | -2.210 | 0.420 | -5.266 | 0.000 | -3.034 | -1.385 |
| Sex: Female | 0.163 | 0.055 | 2.964 | 0.003 | 0.055 | 0.272 |
| Age | 0.046 | 0.005 | 8.906 | 0.000 | 0.036 | 0.057 |
| APOE Status: Non-Carrier | 0.095 | 0.059 | 1.614 | 0.107 | -0.021 | 0.212 |
| Race: African American | 0.046 | 0.119 | 0.387 | 0.699 | -0.188 | 0.280 |
| Race: Asian | -0.224 | 0.128 | -1.750 | 0.081 | -0.475 | 0.028 |
| Race: Hispanic | -0.187 | 0.123 | -1.520 | 0.129 | -0.429 | 0.055 |
| Race: Other or multiple races | -0.138 | 0.110 | -1.253 | 0.211 | -0.355 | 0.079 |
| Education | -0.010 | 0.014 | -0.696 | 0.487 | -0.037 | 0.018 |
| **ADI: Moderate** | **Estimate** | **Std. Error** | **t Value** | **P-value** | **CI Low** | **CI High** |
| Intercept | -2.589 | 0.527 | -4.913 | 0.000 | -3.626 | -1.552 |
| Sex: Female | 0.110 | 0.073 | 1.502 | 0.134 | -0.034 | 0.255 |
| Age | 0.049 | 0.007 | 7.109 | 0.000 | 0.035 | 0.062 |
| APOE Status: Non-Carrier | -0.068 | 0.075 | -0.907 | 0.365 | -0.216 | 0.080 |
| Race: African American | -0.107 | 0.094 | -1.128 | 0.260 | -0.292 | 0.079 |
| Race: Asian | -0.017 | 0.271 | -0.061 | 0.952 | -0.551 | 0.518 |
| Race: Hispanic | -0.017 | 0.120 | -0.140 | 0.889 | -0.252 | 0.219 |
| Race: Other or multiple races | -0.058 | 0.135 | -0.430 | 0.667 | -0.324 | 0.208 |
| Education | 0.012 | 0.016 | 0.765 | 0.445 | -0.019 | 0.043 |
| **ADI: High** | **Estimate** | **Std. Error** | **t Value** | **P-value** | **CI Low** | **CI High** |
| Intercept | -1.343 | 0.990 | -1.356 | 0.180 | -3.320 | 0.634 |
| Sex: Female | 0.107 | 0.161 | 0.665 | 0.509 | -0.215 | 0.429 |
| Age | 0.037 | 0.013 | 2.868 | 0.006 | 0.011 | 0.062 |
| APOE Status: Non-Carrier | -0.296 | 0.147 | -2.021 | 0.047 | -0.589 | -0.004 |
| Race: African American | -0.134 | 0.146 | -0.917 | 0.362 | -0.424 | 0.157 |
| Race: Hispanic | 0.284 | 0.238 | 1.195 | 0.236 | -0.190 | 0.759 |
| Race: Other or multiple races | 0.092 | 0.292 | 0.314 | 0.755 | -0.492 | 0.675 |
| Education | -0.005 | 0.031 | -0.155 | 0.877 | -0.067 | 0.058 |

Abbreviations: APOE = Apolipoprotein E; Std. = Standard; CI = Confidence Interval

Note: Sample size ADI Low = 425; ADI Moderate = 302; ADI High = 73.

**Supplementary Table 34.** Statistics for stratified models by race and ethnicity on WMH. ADI continuous.

| **African American** | **Estimate** | **Std. Error** | **t Value** | **P-value** | **CI Low** | **CI High** |
| --- | --- | --- | --- | --- | --- | --- |
| Intercept | -2.949 | 0.743 | -3.966 | 0.000 | -4.423 | -1.475 |
| Area Deprivation Index. Nation | -0.003 | 0.002 | -1.442 | 0.152 | -0.008 | 0.001 |
| Sex: Female | 0.104 | 0.131 | 0.790 | 0.431 | -0.157 | 0.364 |
| Age | 0.059 | 0.011 | 5.441 | 0.000 | 0.037 | 0.080 |
| APOE Status: Non-Carrier | -0.110 | 0.108 | -1.022 | 0.309 | -0.323 | 0.103 |
| **Asian** | **Estimate** | **Std. Error** | **t Value** | **P-value** | **CI Low** | **CI High** |
| Intercept | -2.986 | 1.205 | -2.479 | 0.021 | -5.485 | -0.488 |
| Area Deprivation Index. Nation | 0.013 | 0.008 | 1.649 | 0.113 | -0.003 | 0.029 |
| Sex: Female | -0.039 | 0.190 | -0.206 | 0.838 | -0.433 | 0.355 |
| Age | 0.048 | 0.018 | 2.596 | 0.017 | 0.010 | 0.086 |
| APOE Status: Non-Carrier | 0.308 | 0.233 | 1.322 | 0.200 | -0.175 | 0.792 |
| **Hispanic** | **Estimate** | **Std. Error** | **t Value** | **P-value** | **CI Low** | **CI High** |
| Intercept | -1.108 | 1.062 | -1.043 | 0.301 | -3.236 | 1.020 |
| Area Deprivation Index. Nation | 0.007 | 0.004 | 1.842 | 0.071 | -0.001 | 0.014 |
| Sex: Female | 0.095 | 0.165 | 0.575 | 0.568 | -0.236 | 0.426 |
| Age | 0.024 | 0.016 | 1.469 | 0.148 | -0.009 | 0.056 |
| APOE Status: Non-Carrier | 0.034 | 0.184 | 0.184 | 0.855 | -0.334 | 0.402 |
| **Other or multiple** | **Estimate** | **Std. Error** | **t Value** | **P-value** | **CI Low** | **CI High** |
| Intercept | -1.141 | 0.878 | -1.301 | 0.199 | -2.903 | 0.620 |
| Area Deprivation Index. Nation | 0.006 | 0.003 | 1.973 | 0.054 | -0.000 | 0.013 |
| Sex: Female | -0.146 | 0.143 | -1.026 | 0.310 | -0.433 | 0.140 |
| Age | 0.028 | 0.012 | 2.332 | 0.024 | 0.004 | 0.053 |
| APOE Status: Non-Carrier | -0.067 | 0.143 | -0.469 | 0.641 | -0.354 | 0.220 |
| **White** | **Estimate** | **Std. Error** | **t Value** | **P-value** | **CI Low** | **CI High** |
| Intercept | -2.446 | 0.335 | -7.294 | 0.000 | -3.104 | -1.787 |
| Area Deprivation Index. Nation | -0.000 | 0.001 | -0.289 | 0.773 | -0.003 | 0.002 |
| Sex: Female | 0.188 | 0.051 | 3.710 | 0.000 | 0.088 | 0.287 |
| Age | 0.048 | 0.005 | 10.010 | 0.000 | 0.039 | 0.057 |
| APOE Status: Non-Carrier | 0.025 | 0.054 | 0.462 | 0.644 | -0.082 | 0.132 |

### **Supplementary Table 35.** Statistics for stratified models by race and ethnicity on WMH. ADI continuous. Education included in model

| **African American** | **Estimate** | **Std. Error** | **t Value** | **P-value** | **CI Low** | **CI High** |
| --- | --- | --- | --- | --- | --- | --- |
| Intercept | -2.609 | 0.872 | -2.990 | 0.003 | -4.338 | -0.879 |
| Area Deprivation Index. Nation | -0.004 | 0.002 | -1.567 | 0.120 | -0.008 | 0.001 |
| Sex: Female | 0.119 | 0.133 | 0.894 | 0.373 | -0.145 | 0.383 |
| Age | 0.059 | 0.011 | 5.426 | 0.000 | 0.037 | 0.080 |
| APOE Status: Non-Carrier | -0.103 | 0.108 | -0.950 | 0.344 | -0.318 | 0.112 |
| Education | -0.021 | 0.028 | -0.749 | 0.455 | -0.077 | 0.035 |
| **Asian** | **Estimate** | **Std. Error** | **t Value** | **P-value** | **CI Low** | **CI High** |
| Intercept | -2.099 | 1.445 | -1.453 | 0.161 | -5.105 | 0.906 |
| Area Deprivation Index. Nation | 0.011 | 0.008 | 1.437 | 0.165 | -0.005 | 0.028 |
| Sex: Female | -0.114 | 0.201 | -0.567 | 0.577 | -0.532 | 0.304 |
| Age | 0.048 | 0.018 | 2.599 | 0.017 | 0.010 | 0.086 |
| APOE Status: Non-Carrier | 0.383 | 0.242 | 1.584 | 0.128 | -0.120 | 0.886 |
| Education | -0.052 | 0.047 | -1.100 | 0.284 | -0.149 | 0.046 |
| **Hispanic** | **Estimate** | **Std. Error** | **t Value** | **P-value** | **CI Low** | **CI High** |
| Intercept | -2.288 | 1.170 | -1.956 | 0.056 | -4.632 | 0.057 |
| Area Deprivation Index. Nation | 0.012 | 0.004 | 2.773 | 0.008 | 0.003 | 0.021 |
| Sex: Female | 0.053 | 0.162 | 0.328 | 0.744 | -0.271 | 0.377 |
| Age | 0.018 | 0.016 | 1.148 | 0.256 | -0.014 | 0.050 |
| APOE Status: Non-Carrier | 0.042 | 0.178 | 0.238 | 0.813 | -0.315 | 0.399 |
| Education | 0.088 | 0.041 | 2.128 | 0.038 | 0.005 | 0.170 |
| **Other or multiple** | **Estimate** | **Std. Error** | **t Value** | **P-value** | **CI Low** | **CI High** |
| Intercept | -0.420 | 1.064 | -0.395 | 0.695 | -2.556 | 1.717 |
| Area Deprivation Index. Nation | 0.006 | 0.003 | 1.691 | 0.097 | -0.001 | 0.012 |
| Sex: Female | -0.151 | 0.142 | -1.064 | 0.293 | -0.437 | 0.134 |
| Age | 0.028 | 0.012 | 2.297 | 0.026 | 0.004 | 0.052 |
| APOE Status: Non-Carrier | -0.051 | 0.143 | -0.358 | 0.722 | -0.338 | 0.236 |
| Education | -0.040 | 0.033 | -1.190 | 0.240 | -0.107 | 0.027 |
| **White** | **Estimate** | **Std. Error** | **t Value** | **P-value** | **CI Low** | **CI High** |
| Intercept | -2.502 | 0.379 | -6.601 | 0.000 | -3.246 | -1.757 |
| Area Deprivation Index. Nation | -0.000 | 0.001 | -0.225 | 0.822 | -0.003 | 0.002 |
| Sex: Female | 0.189 | 0.051 | 3.718 | 0.000 | 0.089 | 0.288 |
| Age | 0.048 | 0.005 | 9.928 | 0.000 | 0.038 | 0.057 |
| APOE Status: Non-Carrier | 0.026 | 0.054 | 0.471 | 0.638 | -0.081 | 0.132 |
| Education | 0.004 | 0.012 | 0.319 | 0.750 | -0.020 | 0.027 |

### **Supplementary Table 36.** Statistics for interaction model. Outcome: gray matter volume. ADI as continuous.

| **term** | **Sum of Squares** | **Degrees of Freedom** | **F value** | **P-value** |
| --- | --- | --- | --- | --- |
| Intercept | 85,672,957.109 | 1 | 133.302 | 0.000 |
| Age | 87,805,656.217 | 1 | 136.620 | 0.000 |
| Area Deprivation Index | 1,058,056.081 | 1 | 1.646 | 0.200 |
| Race | 7,616,624.379 | 4 | 2.963 | 0.019 |
| Sex | 17,982.368 | 1 | 0.028 | 0.867 |
| APOE Status | 447,716.347 | 1 | 0.697 | 0.404 |
| TIV | 670,004,824.116 | 1 | 1,042.488 | 0.000 |
| Area Deprivation Index*Race | 2,999,342.086 | 4 | 1.167 | 0.324 |
| Area Deprivation Index*Sex | 317,706.955 | 1 | 0.494 | 0.482 |
| Area Deprivation Index*APOE Status | 226,624.224 | 1 | 0.353 | 0.553 |
| Residuals | 527,012,345.605 | 820 |  |  |

Abbreviations: APOE = Apolipoprotein E;

Note: Sample size N = 8

### **Supplementary Table 37.** Statistics for interaction model. Outcome: gray matter volume. ADI as categorical

| **term** | **Sum of Squares** | **Degrees of Freedom** | **F value** | **P-value** |
| --- | --- | --- | --- | --- |
| Age | 86,620,933.685 | 1 | 134.725 | 0.000 |
| Area Deprivation Index | 696,626.184 | 2 | 0.542 | 0.582 |
| Race | 10,838,218.493 | 4 | 4.214 | 0.002 |
| Sex | 57,956.866 | 1 | 0.090 | 0.764 |
| APOE Status | 55,308.499 | 1 | 0.086 | 0.769 |
| TIV | 661,408,196.425 | 1 | 1,028.713 | 0.000 |
| Area Deprivation Index*Race | 5,545,562.082 | 7 | 1.232 | 0.282 |
| Area Deprivation Index*Sex | 960,860.604 | 2 | 0.747 | 0.474 |
| Area Deprivation Index*APOE Status | 105,128.965 | 2 | 0.082 | 0.922 |
| Residuals | 523,358,883.617 | 814 |  |  |

Abbreviations: APOE = Apolipoprotein E;

Note: Sample size N =

### **Supplementary Table 38.** Correction for multiple comparisons

|  | CONTINUOUS | | CATEGORICAL | |
| --- | --- | --- | --- | --- |
| Test | Raw p-value | FDR p-value | Raw p-value | FDR p-value |
| Cognition. Sex | 0,701 | 0,770 | 0,417 | 0,592 |
| Cognition. APOE | 0,391 | 0,651 | 0,614 | 0,708 |
| Cognition. Race | 0,057 | 0,425 | 0,04 | 0,150 |
| Amyloid. Sex | 0,32 | 0,607 | 0,46 | 0,592 |
| Amyloid. APOE | 0,265 | 0,607 | 0,001 | 0,015 |
| Amyloid. Race | 0,77 | 0,770 | 0,433 | 0,592 |
| Tau. Sex | 0,732 | 0,770 | 0,015 | 0,112 |
| Tau. APOE | 0,121 | 0,441 | 0,061 | 0,183 |
| Tau. Race | 0,147 | 0,441 | 0,455 | 0,592 |
| WMH. Sex | 0,628 | 0,770 | 0,807 | 0,864 |
| WMH. APOE | 0,014 | 0,210 | 0,025 | 0,125 |
| WMH. Race | 0,085 | 0,425 | 0,465 | 0,592 |
| GM. Sex | 0,482 | 0,723 | 0,474 | 0,592 |
| GM. APOE | 0,553 | 0,754 | 0,922 | 0,922 |
| GM. Race | 0,324 | 0,607 | 0,282 | 0,592 |

### **Supplementary Table 39.** U.S. POINTER study group at baseline

Clinical sites.

North Carolina – Wake Forest University School of Medicine: Jo Cleveland, MD (PI); Jeff

Williamson, MD, MHS (Co-PI); Leslie Teague, MS; Justin Johnson, MS ; Mackenzie Anderson; Philicia

Armstrong, MSW; Derrick Barnes, Margaret Brown; Sarah Brown; Elizabeth Chmelo, MS; Tiffany

Cummings, PsyD (deceased); Elizabeth Dahl; Carlo Davids, MS; Rebecca Davis; Abbie Eaton, PhD,

MMS; Mary Ellenburg; Ana Glover; Anallely Hernandez Laguna; Carolyn Higgins; Christiana Higgs,

MS; Benjamin Hutchison; Ansley Jewell; Rachel Kiger; Michelle Lewis; Kristy Lievense; Marie Liquete,

MS; Kristi Long; Beth Lovette; Brittney McDermott; Melisa Ramirez-Pineda; Andrea Rivis, MS; Cristian

Rivis; Sam Rogers, PA-C; Bonnie Sachs, PhD; Krissi Shook; Lindsay Tysinger; Eileen Weston; Malcom

Williams; Kelvin L. Williams, PhD, D.Min; Dixie Yow; Ezequiel Zamora, MD; Christine Zecca.

Northern California – University of California, Davis: Rachel Whitmer, PhD; Sarah Farias, PhD;

Oanh Meyer, PhD; Anna Garzon, MHA; Kellie Holley; Hollie Adams; Raquel Alto; Ashley Balley,

MPH; Carmen Benavides; Jennifer Cartan, MA; Casey Castro; Manesy Ceja Cevallos; Michelle Chan,

PhD; Carolina Chernyetsky; Evelyn Cordero; Fawn Cothran; Jennifer DeGuzman; Mayra Diaz; Jessica

Famula; Alice Fisher; Marisela Flores; Martha Forloines, PhD; Jessica Geltz; Parvaneh Gerami, FNP;

Jessica Holscher; Jorge Hurtado; Emily Kostner; Elisa Lee; Edward Lingayo Jr; Tyler McConnell;

Macaria Mendoza, MS; Tara Miskovich; Elias Ortiz; Amber Pippins; Magaly Quinteros; Roberto Ramos;

Sarina Rodriguez; Ashley Romo; Lindsay Ruiz Graham; Sandra Ruiz-White; Vanessa Sanchez; Ellen

Thomas; Erica Tutuwan; Itzel Vargas; Hillary Vossler; Kelly Wallace; Derron Yu.

Chicagoland – Rush Colleges of Health Sciences and of Medicine: Christy Tangney, PhD; Martha

Clare Morris, ScD*; Neelum Aggarwal, MD; Meera Sotor, MPH; Kristie Miller; Elizabeth Arrvizu;

Melanie Chavin, MS; Miriam De la Torre; Mindy Dershem, MS; Pankaja Desai, PhD; Sarah Ehlers,

MBA; Bethany Franz, MHA; Tiffini Funches; Jazmin Garcia; Crystal Glover, PhD; Sarah Graef, DC;

Thomas M. Holland, MD; Leah Johnston; Kaitlin Koncilja; Kristin Krueger, PhD; Heidi Langdon, MS;

Brittany Mabry; Mercedes Maceyras; Daniel Madock; Mia McClintic; Miguel Montero; Melissa MoralesPerez, MS; Brooke Nanni, MPH; Radhika Patel, MS; Cristina Quiroz; Nancy Rainwater, MBA; Terrianne

Reynolds, MPH; Chartay Robinson; Jennifer Ventrelle, MS; Genesis Villalobos; Michelle Villanueva,

MS.

Chicagoland – Advocate Health: Darren Gitelman, MD; Masun Jackson, MS; Zeyad Alaswad, MD;

Claudia Amador; Carlos Corado; Maha Haroon; Elizabeth Hartman, PhD; Megon Holldorf; Margaret

Konieczny, MSN; Chauncey Lawson-Weinert; Viet Le; Grace Lucenti; Anthony McCormack, MD;

Elizabeth Omotoye, MPH; Olivia Preissle; Tawny Pyszka; Mary Schmidt; Samuel Streeter; Evelyn

Torres; Sherri Velez; Kathryn Waitzman, MEd.

Houston – Baylor College of Medicine and Kelsey Research Foundation: Valory Pavlik, PhD;

Melissa Yu, MD; Ashley Alexander, MHSA; Michele York, PhD; Sydney O’Connor, MA; Rose TrevinoWhitaker, MPH; Ruchi Aggarwal, MD; Sayo Awosika-Olumo, MD, PhD; Talitha Baszile; Jeffrey

Bishop, PA-C; Regan Brooks; Maricela Caceres; Vanessa Cardenas; Maria Chaudhary, MS; Valerie

Coffman; Jackielynn Cruz; Edina Dervisefendic; Anna Duron; Jacob Faircloth; Jennifer Garrett; Denise

Gibson; Cesar Gonzalez; Latrel Grant; Beck Hill; Kristin Ijeh, MS; Nallely Infante; Chi-Ying Lin, MD,

MPH; Elizabeth Lipscomb, RN; Leah Logan; Erica Lonquich; Patricia Lynch; Bobby Marker; Crystal

Martinez-Busarow; Milena Olivar; Nat Pacini, MA; Arely Perez, MS; Courtney Rice, DNP; Monica

Rodriguear, MA; Demetrio Selman; Kaila Sevilla; Akash Shah; Hannah Shields; Raven Small; Jackie

Soto; Juan Toledo, MD, PhD; John Valenta, PhD; Shayla Yonce.

New England-Rhode Island – Butler Hospital and Miriam Hospital: Stephen Salloway, MD; Stephen

Correia, PhD; Rena Wing, PhD; Meghan Riddle, MD; Tyler Rosenholm; Samuel Slezak, MS; Kathryn

Demos, PhD; KayLoni Olson, PhD; Kelsey Adams; Roseurys Almonte Nova; Nicole Amichetti; Kirsten

Annis; Kathryn Beaulieu; Sarah Benjamin, MS; Sarah Bica; Joni Bloom; Courtney Bodge, PhD; Gregory

Brunson; Brian Castelluccio, PhD; Monique Coley, PA; Karleen Coppola; Ashleigh Cregan; Katherine

Daneault; Linda Davidge; Brittany Dawson, MS; Sarah Deforest; Lyndsay DeMatteo, MSN, Elizabeth

DiGregorio; Caitlin Egan, MS; Sara Eksuzian; Sheina Emrani, PhD; Melanie Faust, MS, FNP-C; Joslynn

Faustino; Angel Garcia; Daisy Garcia; Tiffany Giampa; Brooke Huemann; Shauna Hyde, MS; Athena

Lavoie; Athene Lee, PhD; Heather Maloney; Bill Menard; Andrea Nanos, MSN; Greg Pappas; Bryanne

Peets; Dominique Popescu; Erin Poyant; Ariana Rafanelli; Vivian Ramos; Eliza Rego; Corinne Roma;

Lorrance Saraiva; Sydney Saunders; Tara Tang; Gina Tonini; Priscilla Villa, MS; James Weir.

Coordinating Center.

Administrative and Clinical Operations Coordination Center: Laura Baker, PhD; Nancy Woolard;

Sharon Wilmoth; Amber Adkins Thro; Rebecca Badgio, MS; Brad Caudle; Taylor Dannemiller; Leslie

Gordineer; Allison Heinrich, MS; Marcus Hill, MPH; Cara Johnson, MA; Jeffrey Katula, PhD; Katelyn

King; Desiree Lopez, MSW; Kate Papp, PhD; Margaret Scales, MA; Wilson Sommerville, PhD;

Benjamin Williams, MD, PhD.

Data Coordination Center: Mark Espeland, PhD; Iris Leng, MD, PhD; Laura Lovato, MS; Julia Spell;

Scott Rushing; Debbie Felton; Megan Adkisson; Julissa Almonte; Bobby Amoroso; Ryan Barnard, MS;

Daniel Beavers, PhD; Shyh-Huei Chen, PhD; Danielle Cunio; Yitbarek Demesie, MS; Katie Garcia, MS;

Sarah Gaussoin, MS; KaShawna Guy, MPH; Darrin Harris; Marjorie Howard, MS; Christopher McLouth,

PhD; John Nichols; Jing Su, PhD; Jennifer Talton, MS; Jennifer Walker; Jack White; James Willard,

MAS.

Alzheimer’s Association.

Maria Carrillo, PhD; Heather Snyder, PhD; Susan Antkowiak; Claire Day; Richard Elbein; Ann Marie

McDonald, MBA, MEd; Terrianne Reynolds, MPH; Carl Hill, PhD; Courtney Kloske, PhD; Katherine

Lambert; Heidi Langdon, MS; Olivia Matongo; Emily Meyers, PhD; Joanne Pike, DrPH; April Ross,

PhD; Rebecca Edelmayer, PhD.

Karolinska Institute. Miia Kivipelto, MD, PhD; Tiia Ngandu, MD, PhD; Alina Solomon, MD, PhD.

University of Southern California, Alzheimer’s Therapeutic Research Institute. Robert Rissman, PhD;

Sara Abdel-Latif; Louise Monte, MS; Rema Ramen, PhD.

Vanderbilt University Medical Center. Consuela Wilkins, MD; Tiffany Israel, MSSW.

University of Wisconsin – Madison. Gina Green Harris, MS, PhD.

Posit Science Corporation. Mouna Attarha, PhD; Henry W. Mahncke, PhD.

*Deceased
